# Predicting Intentional Self-Harm Following Psychiatric Discharge in Catalonia, Spain: Machine Learning Models from Linked Registry Data

**DOI:** 10.1101/2025.09.26.25336360

**Authors:** Itxaso Alayo, Oriol Pujol, Franco Amigo, Laura Ballester, Roser Cirici Amell, Salvatore Fabrizio Contaldo, Montserrat Ferrer, Daniel Guinart, Laura Latorre Moreno, Angela Leis, Montserrat Lopez Fernandez, Miguel Angel Mayer, Manuel Pastor, Carlos Peña-Salazar, Ana Portillo-Van Diest, Juan Manuel Ramírez-Anguita, Ferran Sanz, Jordi Alonso, Ronald C Kessler, Lars Mehlum, Diego Palao, Víctor Pérez Sola, Gemma Vilagut, Philippe Mortier

## Abstract

**Introduction:** Patients recently discharged from psychiatric hospitalization are at increased risk of intentional self-harm, including suicide. Using linked population-based registry data from Catalonia, Spain, we developed machine learning-based prediction models for post-discharge intentional self-harm across different follow-up horizons, sex, and age groups, and evaluated their generalizability and robustness with multiple validation strategies.

**Methods:** Retrospective cohort study including 41,827 individuals accounting for 71,865 psychiatric hospitalizations with discharge at age ≥10 years, between January 1, 2015, and December 31, 2018, in Catalonia, Spain, with follow-up until December 31, 2019. Primary outcome was intentional self-harm (fatal or non-fatal) within 7, 30, 90, 180, and 365 days post-discharge. Models incorporated 247 predictors from electronic health records, including sociodemographic characteristics, mental and physical disorder categories, categories of dispensed psychotropic medication, and history of self-harm and psychiatric hospitalization. Model performance was evaluated using the area under the receiver operating characteristic curve (AUCROC) and the area under the precision-recall curve (AUCPR). Predictor importance was assessed using Shapley Additive Explanations (SHAP).

**Results:** Within 365 days, 4,901 hospitalizations (6.8%) were followed by intentional self-harm. The 365-day model trained on the full cohort achieved a AUCROC of 0.819, in the test sample with adjusted AUCPR indicating a median 5.4-fold improvement over baseline prevalence. This model generalized well across event horizons and sex–age strata, outperforming subgroup-specific models when data sparsity limited performance. Separate models trained by event horizons, and stratified by sex, and sex–age groups achieved a median AUCROC of 0.775 (IQR 0.764–0.808), with adjusted AUCPR indicating a median 5.4-fold improvement over baseline prevalence (IQR 4.5–6.2). Key predictors included the recency of the last registered diagnosis of depressive episodes, recurrent depression, adjustment disorders, and schizophrenia, as well as recent SSRI dispensation and the number of childhood-onset disorder and musculoskeletal disease diagnoses in the previous five years. Predictor importance varied considerably across sex–age strata, with smaller differences across horizons. Subject-level and temporal split validation strategies reduced performance (AUCROC 0.711–0.746), though estimates remained clinically informative (2.8–3.1-fold improvement over baseline prevalence).

**Conclusions:** Machine learning models using routinely collected health records predicted intentional self-harm after psychiatric hospitalization with good discrimination and clinically meaningful precision–recall performance. A single 365-day model generalized well across horizons and demographic groups, suggesting that one broadly trained model may provide a pragmatic and scalable approach for clinical implementation.

## INTRODUCTION

Self-harm is a major public health concern [1–3], with about 5.5 million incident cases worldwide each year; this annual incidence is projected to be 10.6 million by 2040 [4]. Self-harm ranks among the leading causes of mortality and morbidity worldwide [5], particularly among individuals with psychiatric disorders [6]. The assessment and management of patients with risk for self-harm represents a significant clinical challenge [7,8]. One subgroup with high risk for self-harm are patients recently admitted in a psychiatric ward, with the period immediately following discharge from psychiatric hospitalization being associated with a markedly elevated risk of behavior compared to the general population, particularly during the first few weeks post-discharge [9–12]. In Catalonia, Spain, it has been estimated that patients with a recent psychiatric hospitalization are at a markedly elevated risk of both nonlethal intentional self-harm (NLISH) and suicide, with 5-year cumulative incidence of NLISH being 14.2% for females and 8.7% for males, and standardized mortality rates for suicide being 47.6 and 27.9, respectively [11]. This highlights the need for effective risk assessment strategies to guide preventive interventions tailored to this high-risk clinical setting. However, both clinical judgment and risk assessment scales lack the necessary predictive validity for suicide and non-lethal self-harm [13,14]. Therefore, novel predictive tools that can more reliably identify individuals at imminent risk and guide targeted prevention in the post-discharge setting are needed.

In this context, machine learning (ML) has emerged as a promising alternative to traditional statistical methods for developing clinical prediction models for self-harm [15,16], particularly given the increasing availability of electronic health records (EHR), which capture data from large and clinically representative populations [17–19]. A recent systematic review and meta-analysis of ML models for suicide-related outcomes reported a pooled area under the curve (AUC) of 0.86, with a sensitivity of 0.66 and specificity of 0.87, highlighting their potential to inform real-world clinical decision-making [20]. Unlike conventional approaches, ML models can flexibly incorporate a wide range of potentially relevant predictors, including sociodemographic characteristics, mental and somatic disorder diagnoses and medication use, while also accounting for the temporal relationships between these predictors, as reflected in the structure of EHR data [21]. Including temporal information on mental and physical disorder diagnosis is particularly valuable, as models that incorporate time-sensitive information have consistently outperformed those that rely solely on static features [22,23].

Despite these promising developments, several important knowledge gaps remain to enable real-time implementation of self-harm prediction models. First, few studies have specifically developed ML-based models for self-harm following psychiatric hospital discharge. Most have focused primarily on suicide as the outcome [24–27], and, to our knowledge, only one has also included non-lethal intentional self-harm [26]. Second, relatively few studies have developed separate models by sex [28], age group [20], or prediction horizon [25,29], nor systematically identified whether the most relevant predictors differ across such stratified models. Although sex [30,31] and age [32] are well-established modifiers of suicidal behavior, most existing work has not explicitly examined how model performance or predictor importance varies across these subgroups [33] or across short-versus long-term outcomes. Third, most previous studies have instead focused on general models trained for a single, typically short-term prediction horizon (e.g., 30- or 90-day risk) [26,34]. Far fewer have evaluated whether such models generalize robustly across multiple temporal windows of risk or across demographic subgroups such as sex and age [35–37]. Fourth, there is limited understanding of how different data splitting strategies (e.g., episode-level, subject-level, or temporal splits) affect model performance, validity and clinical applicability of predictive models [38,39]. In clinical practice, the repeated hospitalizations of the same patient provide valuable information for risk assessment. However, it is unclear to what extent models benefit from historical episodes versus information from new patients. It is essential to evaluate these different strategies to ensure that predictive tools perform accurately for both; new and returning patients, and that they can be reliably applied in real-world clinical settings where patients may be admitted repeatedly [40].

The present study has three objectives: (1) to develop machine learning-based prediction models for intentional self-harm following discharge from psychiatric hospitalization using electronic health record data, evaluate their predictive accuracy, and to examine whether the relative importance of predictors and model performance varies by follow-up period (7 to 365 days), sex, and age group; (2) to assess the generalizability of a 365-day prediction model trained on the full cohort (both sexes, all ages) to shorter follow-up periods and to specific sex and age groups; and (3) to evaluate the impact of different data splitting strategies (episode-level splits, subject-level splits, random single-episode selection per patient, temporal splits) on robustness and generalizability of the models.

## METHODS

### Study Population and Design

This is an observational retrospective cohort study including all patients with psychiatric hospitalizations in Catalonia, Spain (total population 7.6 million) between 1 January 2015 and 31 December 2018. The sample comprised all hospitalization episodes of the study cohort during this period, limited to those in which individuals were aged 10 years or older at the time of discharge. Sample follow-up was extended to 31 December 2019 for outcome assessment (i.e., self-harm, defined as either NLISH or suicide) to ensure that each hospitalization had a retrospective follow-up period of at least 12 months.

The research protocol received approval from the Parc de Salut Mar clinical research ethics committee (CEIC protocol 2017/7431/I), which granted a waiver for individual informed consent due to the fully anonymized nature of all registry data and comprehensive reidentification risk assessment procedures.

### Data Sources and Registry Integration

We obtained registry data from the Agency for Health Quality and Assessment of Catalonia, which coordinates the Public Data Analysis for Health Research and Innovation Program [41]. Individual records were linked across multiple registries using personal healthcare identification numbers. The linked datasets comprised: (1) mortality records from the Spanish National Statistics Institute (2015–2019) [42], including suicide deaths identified through International Classification of Diseases, Tenth Revision (ICD-10) codes [43]; (2) routinely collected electronic health records (EHR) from the Catalan Health Service [44], covering the entire public healthcare network and including psychiatric hospital admissions (2008–2019), outpatient mental health visits (2008–2019), emergency department contacts (2014–2019), general hospital admissions (2007–2019), and primary care visits (2010–2019), with diagnoses coded using ICD-9-CM, ICD-10, or ICD-10-CM [45–47]. The Catalan Health Department ensures EHR data quality through systematic verification and validation by trained health documentalists; (3) records of psychotropic medication dispensations by community pharmacies (2014–2019), coded according to the Anatomical Therapeutic Chemical (ATC) classification system [48,49]; (4) data on clinically confirmed self-harm episodes (2014–2019) reported through the Catalonia Suicide Risk Code (CSRC) program [50], a region-wide suicide prevention protocol that mandates in-person psychiatric assessment for residents presenting with self-harm or suicide risk in any public healthcare setting; and (5) administrative data from the Catalan Health Service’s central population register (2014–2019), including demographic variables such as sex, age, socioeconomic status, and nationality.

### Study outcome

The primary outcome was intentional self-harm, either lethal (suicide; identified using ICD-10 codes X60–X84) or non-lethal intentional self-harm (NLISH, ascertained through two complementary approaches: i) healthcare encounters coded with standard external cause codes for self-harm in routine EHR data (ICD-9-CM E950–E959, ICD-10 X60*–X84* and Y87.0, or ICD-10-CM T14.91*, X71*–X83*, Y87.0, and T36*–T65*/T71* when the fifth or sixth character indicated ‘2’), and ii) episodes documented in the CSRC program with a clinically verified self-harm method. The first recorded NLISH episode following each psychiatric hospitalization, captured in either source, was used to define the event.

The outcome was assessed at the level of each psychiatric hospitalization episode. Outcome was calculated for five predefined temporal horizons: 7, 30, 90, 180, and 365 days post-discharge, allowing for both short- and long-term risk prediction models.

### Predictor variables

For each psychiatric hospitalization, we constructed the following predictor variables, divided into 5 classes: (1) sociodemographic characteristics, including biological sex (male/female) and age at discharge (continuous); (2) 51 mental disorder categories created by grouping related mental disorder ICD codes (see **Supplementary Table 1** for details) recorded in the previous five years, (3) 10 physical disorder categories created by grouping associated ICD codes (see **Supplementary Table 1**) recorded in the previous five years; (4) 28 psychotropic medication categories created by grouping ATC codes (see **Supplementary Table 1**) of psychotropic medications dispensed within the past year; and (5) number of prior psychiatric hospitalizations in the past five years; and (6) NLISH diagnoses, recorded in the past year at either the emergency department or through the CSRC program. Mental and physical disorder diagnoses linked to emergency department visits were extracted for the preceding year only, as such information was available from 2014 onwards. Similarly, since both emergency department data and CSRC program data were available as from 2014, prior NLISH diagnoses were extracted for the preceding year only. Since psychotropic medication (ATC codes) dispensing dates were available only at the month–year level, we set them to the first day of the subsequent month to prevent creating predictors that reflect medication dispensed after the index hospitalization (i.e., to avoid post-hospitalization information leakage).

For all mental and physical disorder diagnoses, and for NLISH diagnoses, we derived three types of predictor variables: (1) current diagnosis, a binary indicator of whether the diagnosis was recorded for the current (index) psychiatric hospitalization; (2) number of prior diagnoses, defined as the total number of times the diagnosis was recorded in the previous five years (1 year for NLISH diagnoses) across any healthcare setting, excluding diagnoses associated with the current psychiatric hospitalization; (3) recency of last registered diagnosis, calculated for diagnoses in the previous five years (1 year for NLISH diagnoses), again excluding diagnoses associated with the current psychiatric hospitalization, using the formula exp(–0.02 × [(date of current hospitalization – date of most recent diagnosis) / 30]), resulting in a continuous variable ranging from 0 to 1, with higher values indicating diagnoses closer in time to the current (index) hospitalization. For individuals without prior diagnoses in the previous five years, the recency variable was set to 0. In the case of medications, the variables number of prior psychotropic medications and recency of psychotropic medications are created in the same way as the diagnoses. In addition, for each psychotropic medication, the variable current psychotropic medication was constructed as a binary indicator of whether that psychotropic medication had been dispensed to the patient during the past year.

A total of 247 variables were included in the model. All predictive variables were constructed using information available during the index episode and up to that point, avoiding post-hospitalization information leakage of data on future outcomes.

### Statistical analysis

First, sample characteristics of hospitalizations were summarized as distribution percentages of principal sociodemographic and outcome variables. All study variables were complete, with no missing data identified.

To address objective 1, separate machine learning models were developed to predict intentional self-harm across multiple temporal horizons (7, 30, 90, 180, and 365 days) after discharge from psychiatric hospitalization. For the 365 days temporal horizon, models were also fitted stratified by sex to account for potential sex-specific risk factors and patterns in intentional self-harm. Additional stratification was performed by dividing patients into three age categories (less than 25, 25 -64, more than 64) within each sex group to enable age-specific risk assessment within sex groups and understanding of developmental and life-stage influences on intentional self-harm risk. The dataset included all available psychiatric hospitalizations (i.e., both single as well as repeat hospitalizations) and was split into training (80%) and test sets (20%) at the episode (hospitalization) level, reflecting clinical practice, where repeated assessments of the same individual are common. Random stratified sampling was used in the splitting to ensure that the proportion of positive cases was the same in both training and testing datasets. A 5-fold stratified cross-validation (StratifiedKFold) [51] was implemented during the training phase to ensure robust model performance estimation and prevent overfitting. In each fold, FLAML (Fast and Lightweight Automated Machine Learning) [52–54] was used to search for the best model and hyper-parameters among the following models: Light Gradient Boosting Machine (LightGBM) [55], XGBoost [56], Random Forest [57], Extremely Randomized Trees (Extra Trees) [58], K-Nearest Neighbors (KNN) [59] and Logistic Regression (without penalty, with L1 penalty, and with L2 penalty) [60]. AutoML was configured to optimize model performance using the Area Under the Receiver Operating Characteristic Curve (AUCROC), due to its effectiveness in measuring discrimination for binary classification tasks [61]. Finally, the optimal configuration identified by FLAML was retrained on the entire training dataset (including all cross-validation folds). The final model was then applied to the test set to obtain the validation metrics.

Model performance was assessed using multiple evaluation metrics appropriate for binary classification problems. The primary evaluation metric was the area under the precision-recall curve (AUCPR). Although AUCPR is informative for imbalanced datasets [62], its absolute value is highly dependent on the prevalence of the positive cases. To contextualize performance, AUCPR for the different models were adjusted by dividing by the baseline performance of a random classifier, which corresponds to the prevalence of positive cases. This adjustment allows interpretation of AUCPR relative to what could be achieved by chance, indicating whether the model performs better than random guessing [26]. AUCROC curves were also reported as a complementary metric [63]. Confidence intervals at the 95% level for all metrics were estimated using bootstrap resampling.

SHapley Additive exPlanations (SHAP) values were calculated to identify the most important risk factors for intentional self-harm prediction [64]. SHAP analysis provided global feature importance ranking of variables by their average contribution to predictions across all samples and individual prediction explanations for understanding specific risk factors for individual patients. This analysis enabled quantification of each variable’s contribution to the prediction, understanding of directional relationships between variables and intentional self-harm risk.

To evaluate the generalizability of the predictive model developed on the full cohort for the 365-day time horizon (objective 2), we first tested its model performance for predicting the other temporal horizons. This approach aimed to examine whether a single model could provide consistent predictive performance across different temporal outcomes. In addition, model performance was assessed within subgroups defined by sex, and sex and age, to explore potential differences in predictive accuracy across these populations.

To assess objective 3, a complementary analysis was carried out exploring additional data-splitting and selection strategies to assess the stability of the predictive models under different validation schemes. The following approaches were tested: (i) subject-level split, grouping all episodes of the same individual together in either train or test sample [21,65]; (ii) temporal splits, where earlier years were used for training and subsequent years for validation (2015–2017 vs. 2018) [38,66]; and (iii) random selection of a single episode per subject [67,68].

## RESULTS

### Sample characteristics

The study sample comprised 41,827 individuals with a total of 71,865 psychiatric hospitalizations. Of these, 66.2% experienced a single psychiatric hospitalization in the 2015-2018 observation period, 18.5% had two hospitalizations, and 14.7% had three or more (see **Supplementary Table 2**). Males accounted for 52.9% of all hospitalizations; most admissions involved individuals aged 25–64 years (65.9% among females and 76.6% among males; see **Supplementary Table 3**). Among females, 17.1% were aged over 65 years, compared with only 8.9% of males. At 365-day follow-up after discharge, 4,901 hospitalizations (6.8%) were followed by intentional self-harm; among females this was 2,918 (8.6%) and among males 1,983 (5.2%). Of these, 331 cases (6.8%) were lethal (suicides), including 127 among females (4.4%) and 204 among males (10.3%).

The proportion of hospitalizations followed by intentional self-harm increased from 0.7% at 7 days to 6.8% at 365 days of follow-up (see **Supplementary Table 4**). For all event horizons, rates of post-discharge intentional self-harm were higher for females and for those aged <25 years, and lower for those aged ≥65 years. Rates were also higher for those with prior psychiatric hospitalizations and previous NLISH diagnoses, and with NLISH diagnosis associated with the index hospitalization.

### Prediction models by temporal horizons

After evaluating different models, LightGBM achieved the best performance across all prediction horizons, with the exception of the 90-day horizon, where Extra Trees performed best, and the 7-day horizon, where XGBoost was the top-performing model (see **Supplementary Table 5** for further information on best fitting model and hyperparameter values).

Model performance was evaluated across prediction horizons ranging from 7 to 365 days. Discrimination as measured by the AUCROC was consistently high, ranging from 0.764 for the 30-day horizon to 0.819 for 365 days. Adjusted **AUCPR** values indicated that predictive performance improved 5.4 to 7.1-fold over random classification (i.e., baseline prevalence), with the highest relative gains observed for the 7 and 90-day event horizons and the lowest gain for the 365-day horizon (see **Table 1** and **Supplementary Figure 1**).

**Table 1.** Model performance metrics by event horizon, sex, and sex-age group.

|  | Outcome Prevalence | AUCROC<br>(95% CI) | AUCPR<br>(95% CI) | Adjusted AUCPR<br>(95% CI) |
| --- | --- | --- | --- | --- |
| **Event horizons** |  |  |  |  |
| Full sample, 365 days | 6.8 | 0.819 (0.805 - 0.834) | 0.370 (0.338 - 0.401) | 5.4 (5.0 -5.9) |
| Full sample, 180 days | 4.7 | 0.810 (0.792 - 0.827) | 0.288 (0.253 - 0.324) | 6.2 (5.5 -6.9) |
| Full sample, 90 days | 3.2 | 0.806 (0.786 - 0.827) | 0.208 (0.172 - 0.246) | 6.6 (5.5 -7.8) |
| Full sample, 30 days | 1.7 | 0.764 (0.733 - 0.795) | 0.104 (0.071 - 0.144) | 6.2 (4.3 -8.7) |
| Full sample, 7 days | 0.7 | 0.775 (0.726 - 0.824) | 0.050 (0.028 - 0.093) | 7.1 (4.2 -13.5) |
| **Sex** |  |  |  |  |
| Female, 365 days | 8.6 | 0.801 (0.782 - 0.821) | 0.390 (0.352 - 0.433) | 4.5 (4.1 -5.5) |
| Male, 365 days | 5.2 | 0.773 (0.747 - 0.799) | 0.281 (0.234 - 0.326) | 5.4 (4.6 -6.3) |
| **Sex and age** |  |  |  |  |
| Female < 25, 365 days | 15.0 | 0.767 (0.730 - 0.805) | 0.450 (0.377 - 0.527) | 3.0 (2.5 -3.6) |
| Female [25 ,64], 365 days | 8.4 | 0.808 (0.784 - 0.832) | 0.409 (0.358 - 0.458) | 4.9 (4.3 -5.5) |
| Female > 64, 365 days | 3.1 | 0.754 (0.673 - 0.835) | 0.111 (0.054 - 0.236) | 3.6 (2.0 -7.9) |
| Male < 25, 365 days | 4.6 | 0.737 (0.653 - 0.821) | 0.249 (0.141 - 0.400) | 5.4 (3.3 -8.9) |
| Male [25 ,64], 365 days | 5.6 | 0.835 (0.801 - 0.859) | 0.444 (0.390 - 0.498) | 8.0 (7.0 -9.2) |
| Male > 64, 365 days | 3.3 | 0.525 (0.397 - 0.653) | 0.054 (0.021 - 0.177) | 1.6 (0.8 -6.1) |

*Model performance metrics have been evaluated in the test dataset. AUCROC: area under the receiver operating characteristic curve. AUCPR: area under the precision-recall curve. Adjusted AUCPR= AUCPR divided by the prevalence of positive cases.*

Across all temporal horizons, four diagnosis-recency variables consistently ranked among the top predictors: depressive episode, adjustment disorder, mixed anxiety–depression, schizophrenia and borderline personality disorder. Predictors recurring in two or more horizon-specific models included: recency and number of unspecified mental disorder diagnoses; mood-related predictors (recency of SSRI dispensing, recency of recurrent depression and number of prior depression diagnoses); number of mixed anxiety and depression diagnoses; recency of diagnoses of non-borderline personality disorder, unspecified nonorganic psychosis, pain symptoms and musculoskeletal disease; having an external cause related to accidents or homicide associated with the index hospitalization; and number of second-generation antipsychotics dispensed. Regarding medications, the most important predictor at 365 days was the use of SSRIs, while for the remaining temporal horizons except 30-day models, second-generation antipsychotics were the most relevant. Of note, the number of prior self-harm episodes was the second most important predictor in the 90-day horizon model, and recency of self-harm was the third most important in the 180-day horizon model. No other self-harm–related variables ranked as important in the 7-day, 30-day or 365-day horizon models (**Table 2** and **Supplementary Figure 2**).

**Table 2.** Shapley additive explanation (SHAP) values for prediction models by event horizon.

|  | 7days | 30days | 90days | 180days | 365days |
| --- | --- | --- | --- | --- | --- |
| Depressive episode -recency | 0.18835 | 0.00097 | 0.08042 | 0.25596 | 0.30152 |
| Adjustment disorders –recency | 0.05181 | 0.00117 | 0.05483 | 0.12391 | 0.12802 |
| Mixed anxiety-depression –recency | 0.12297 | 0.00076 | 0.04797 | 0.08150 | 0.12664 |
| Borderline PD –recency | 0.02526 | 0.00052 | 0.01942 | 0.07197 | 0.05250 |
| Schizophrenia –recency | 0.06154 | 0.00017 | 0.02611 | 0.10409 | 0.04763 |
| Second-gen antipsychotics -recency | 0.05161 |  | 0.03994 | 0.06817 | 0.07611 |
| Other personality disorders –recency |  |  | 0.01990 | 0.03469 | 0.07169 |
| Unspecified mental disorder –recency |  | 0.00019 |  | 0.07341 | 0.06599 |
| Depressive episode –number* |  | 0.00011 | 0.02557 |  | 0.04877 |
| Mixed anxiety-depression –number | 0.02888 |  | 0.03497 | 0.07229 |  |
| SSRIs –recency |  |  |  | 0.05358 | 0.16466 |
| Recurrent depression –recency |  |  | 0.01931 |  | 0.08429 |
| Unspecified mental disorder –number |  |  |  | 0.03764 | 0.06986 |
| Musculoskeletal diseases –recency |  | 0.00010 |  | 0.05507 |  |
| Unspecified nonorganic psychosis –recency | 0.04604 |  |  | 0.03980 |  |
| Second-gen antipsychotics -number | 0.03513 |  | 0.02513 |  |  |
| Schizophrenia -number | 0.02943 |  | 0.02278 |  |  |
| External causes -yes/no | 0.04340 | 0.00036 |  |  |  |
| Pain symptoms –recency | 0.05632 | 0.00018 |  |  |  |
| Endocrine/metabolic disorders -number |  |  |  |  | 0.11369 |
| Endocrine/metabolic disorders -recency |  |  |  |  | 0.11215 |
| Pain symptoms -number |  |  |  |  | 0.06192 |
| Previous self-harm -recency |  |  |  | 0.11087 |  |
| Anxiolytics -recency |  |  |  | 0.03813 |  |
| Previous self-harm -number |  |  | 0.07165 |  |  |
| Previous urgent hospitalizations -number |  |  | 0.06442 |  |  |
| MH/SUD screening/history -number |  |  | 0.02960 |  |  |
| Persistent mood disorders -recency |  | 0.00028 |  |  |  |
| Adjustment disorders -yes/no |  | 0.00020 |  |  |  |
| Eating disorders -recency |  | 0.00016 |  |  |  |
| Adjustment disorders -number |  | 0.00011 |  |  |  |
| Borderline PD -number |  | 0.00008 |  |  |  |
| Bipolar disorder -recency | 0.03521 |  |  |  |  |
| Circulatory diseases -recency | 0.02835 |  |  |  |  |
| Opioid analgesics -recency | 0.02834 |  |  |  |  |

### Prediction models by sex

The females specific model achieved an AUCROC of 0.801, and an adjusted AUCPR of 4.5. For males, performance was similar, with an AUCROC of 0.773, and an adjusted AUCPR of 5.4. While overall discrimination was nearly identical across sexes, the adjusted AUCPR indicated relatively greater improvement over baseline prevalence in males (See **Table 1**).

Several predictors of recent diagnosis were consistently important across both sexes, including adjustment disorder, depressive episode, schizophrenia-spectrum disorders, mixed anxiety–depression, non-borderline personality disorder and recent SSRI dispensing. The patterns specific to each sex largely reflected those in the 365-day model for the full cohort, with additional sex-specific predictors detailed in **Supplementary Table 6** and **Supplementary Figure 3**.

### Prediction models by sex-age groups

In the analyses stratifying for both sex and age group, model performance showed variability across subgroups (See **Table 1**). Among females, the best performance was in the 25–64 age group (adjusted AUCPR = 4.9). Results were lower and similar in both younger (<25 years; adjusted AUCPR = 3.0) and older females (>64 years; adjusted AUCPR = 3.6). For males, the highest performance was also in the 25–64 group (adjusted AUCPR = 8.0). Performance was lower in younger males (<25 years; adjusted AUCPR = 5.4) and considerably lower in those over 64 (AUCPR = 1.6). Taken together, these findings suggest that predictive performance was strongest in middle-aged groups for both sexes.

Age stratification among females (**Table 3** and **Supplementary Figure 4**) revealed that relatively few predictor variables consistently ranked among the most important across two or more of the three age-specific prediction models. For females aged 25–64 years, the strongest predictors were recency of schizophrenia diagnosis, mixed anxiety–depression, borderline or other personality disorder, depressive episode, and adjustment disorder. Among females aged ≤24 years, the most important predictor was current SSRI dispensation, followed by recency of mixed anxiety–depression diagnosis, number of non-borderline personality disorder diagnoses, recency of childhood-onset disorder diagnosis, current dispensation of second-generation antipsychotics, and adjustment disorder associated with the index hospitalization. In females aged ≥65 years, the leading predictor was recency of recurrent depression, followed by number of respiratory disease diagnoses, recency of second-generation antipsychotic dispensation, and recency of diagnoses for digestive diseases, organic mental disorders, and non-borderline personality disorders.

**Table 3.**
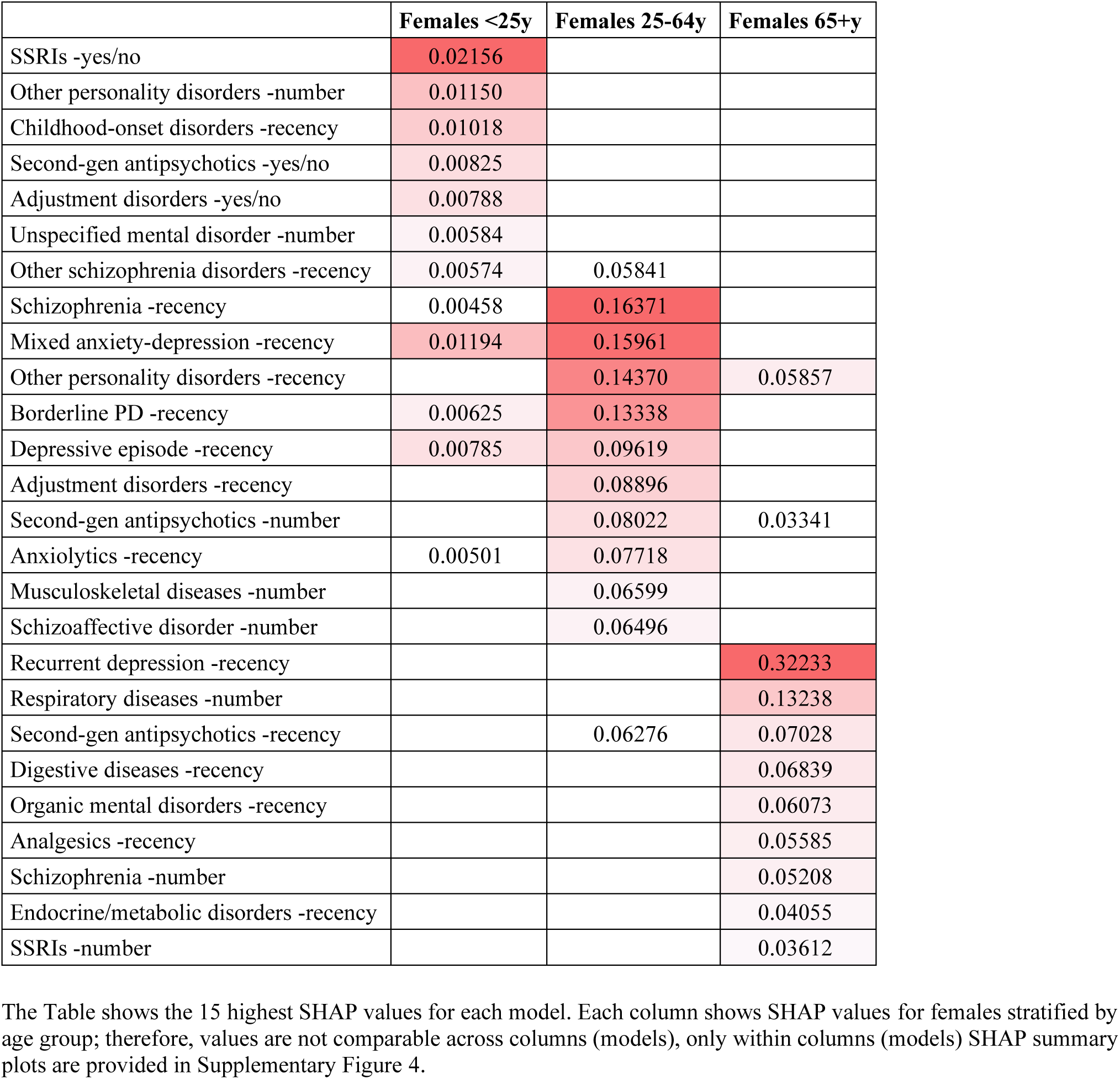
Shapley additive explanation (SHAP) values for prediction models among females stratified by age group.

Age stratification among males (**Table 4** and **Supplementary Figure 5**) revealed that very few predictor variables consistently ranked among the most important across two or more of the three age-specific prediction models. For males aged 25–64 years, the strongest predictors were the number of musculoskeletal disease diagnoses, number of pain-related diagnoses, recency of depressive episode diagnosis, cocaine use disorder, alcohol use disorder, non-borderline personality disorder, unspecified mental disorder, and the number of external cause diagnoses related to accidents and homicide. Among males aged ≤24 years, the most important predictors were the number of childhood-onset disorder diagnoses, recency of schizophrenia diagnosis, the number of positive screens or registered history of mental health and substance use codes, number of endocrine–metabolic disease diagnoses, recency of tobacco use disorder diagnosis, number of cocaine use disorder diagnoses, and recency of registered external cause codes related to accidents or homicide. In males aged ≥65 years, the strongest predictor was recency of depressive episode diagnosis, followed by number of somatoform disorder diagnoses, index hospitalization for non-borderline personality disorder, recency of second-generation antipsychotic dispensation and sedative use disorder diagnosis, and number of second-generation antipsychotic and anxiolytic dispensations.

**Table 4.** Shapley additive explanation (SHAP) values for prediction models among males stratified by age group.

|  | Males <25y | Males 25-64y | Males 65+y |
| --- | --- | --- | --- |
| Childhood-onset disorders -number | 0.12856 |  |  |
| Schizophrenia -recency | 0.10182 | 0.00125 |  |
| MH/SUD screening/history -number | 0.09158 |  |  |
| Endocrine/metabolic disorders -number | 0.08078 |  |  |
| Tobacco use disorder -recency | 0.07596 |  | 0.00072 |
| Cocaine use disorder -number | 0.07387 |  |  |
| External causes -recency | 0.05209 |  |  |
| Developmental disorders -number | 0.05088 |  |  |
| Mood stabilizers -number | 0.04979 |  |  |
| Adjustment disorders -recency | 0.04389 |  |  |
| Mixed anxiety-depression -recency | 0.03561 | 0.00108 |  |
| Musculoskeletal diseases -number |  | 0.01006 |  |
| Pain symptoms -number |  | 0.00435 |  |
| Cocaine use disorder -recency |  | 0.00325 |  |
| Alcohol use disorder -recency |  | 0.00268 |  |
| Other personality disorders -recency |  | 0.00238 |  |
| Unspecified mental disorder -recency |  | 0.00238 |  |
| External causes -number |  | 0.00197 |  |
| Bipolar disorder -number |  | 0.00153 |  |
| SSRIs -recency |  | 0.00139 |  |
| Depressive episode -recency |  | 0.00362 | 0.05462 |
| Somatoform disorders -number |  |  | 0.02485 |
| Other personality disorders -yes/no |  |  | 0.00559 |
| Second-gen antipsychotics -recency |  |  | 0.00473 |
| Sedative use disorder -recency |  |  | 0.00325 |
| Second-gen antipsychotics -number |  |  | 0.00210 |
| Anxiolytics -number |  |  | 0.00151 |
| Cannabis use disorder -number |  |  | 0.00150 |
| Generalized anxiety -number |  |  | 0.00148 |

Detailed SHAP analyses (**Supplementary Figure 4 & 5**) across sex- and age-specific models indicated that several predictors, including mood disorders (depressive episode, recurrent depression), borderline personality disorder, drug use disorders such as cocaine and sedative use disorder, adjustment disorder, and developmental disorders, were almost exclusively associated with increased risk of self-harm within the corresponding subgroups. In contrast, a range of other predictors demonstrated bidirectional contributions to risk, with their effects varying not only across sex and age strata but also within strata depending on co-occurring clinical characteristics captured by the decision tree–based algorithms. For example, schizophrenia-related diagnoses, second-generation antipsychotic dispensations, bipolar disorder, non-borderline personality disorder diagnoses, mixed anxiety–depression, external causes of accidents, alcohol and cannabis use disorder, and childhood disorders all showed context-dependent effects, whereby diagnosis recency or frequency could either increase or attenuate risk depending on the subgroup or additional clinical features.

### Generalizability of 365-day model across time horizons and sex-age subgroups

As shown in **Table 5**, the full-cohort model trained for intentional self-harm at 365-day performed similarly to models trained specifically for each period. Importantly, the adjusted AUCPR values of the 365-day model were consistently similar with those of the horizon-specific models (e.g., 5.8 vs. 6.2 at 180 days, 6.3 vs. 6.2 at 30 days, and 8.2 vs. 7.1 at 7 days), indicating that the full-cohort model maintained equal relative performance.

**Table 5.** Evaluation of the 365-day model’s generalizability across time horizons and sex- and sex-age subgroups.

| Train | Test | Outcome Prevalence | AUCROC (95% CI) | AUCPR (95% CI) | Adjusted AUCPR (95% CI) |
| --- | --- | --- | --- | --- | --- |
| Event horizons |  |  |  |  |  |
| 180 days | Full-cohort, 180 days | 4.7 | 0.810 (0.792 - 0.827) | 0.288 (0.253 - 0.324) | 6.2 (5.5 -6.9) |
| Full-cohort, 365 days |  | 4.7 | 0.823 (0.806 -0.84) | 0.276 (0.241 -0.312) | 5.8 (5.1 -6.6) |
| 90 days | Full-cohort, 90 days | 3.3 | 0.806 (0.786 - 0.827) | 0.208 (0.172 - 0.246) | 6.6 (5.5 -7.8) |
| Full-cohort, 365 days |  | 3.3 | 0.817 (0.797 -0.837) | 0.202 (0.168 -0.24) | 6.2 (5.2 -7.3) |
| 30 days | Full-cohort, 30 days | 1.7 | 0.764 (0.733 - 0.795) | 0.104 (0.071 - 0.144) | 6.2 (4.3 -8.7) |
| Full-cohort, 365 days |  | 1.7 | 0.809 (0.781 -0.837) | 0.105 (0.075 -0.14) | 6.3 (4.6 -8.4) |
| 7 days | Full-cohort, 7 days | 0.7 | 0.775 (0.726 - 0.824) | 0.050 (0.028 - 0.093) | 7.1 (4.2 -13.5) |
| Full-cohort, 365 days |  | 0.7 | 0.795 (0.752 -0.838) | 0.054 (0.026 -0.097) | 8.2 (4.1 -15.1) |
| Sex |  |  |  |  |  |
| Female, 365 days | Female, 365 days | 8.6 | 0.801 (0.782 - 0.821) | 0.390 (0.352 - 0.433) | 4.5 (4.2 -5.0) |
| Full-cohort, 365 days |  | 8.8 | 0.824 (0.806 -0.841) | 0.409 (0.367 -0.45) | 4.6 (4.2 -5.1) |
| Male, 365 days | Male, 365 days | 5.2 | 0.773 (0.747 - 0.799) | 0.281 (0.234 - 0.326) | 5.4 (4.6 -6.3) |
| Full-cohort; 365 days |  | 5 | 0.797 (0.772 -0.822) | 0.302 (0.255 -0.352) | 6.0 (5.1 -7.0) |
| Sex and age |  |  |  |  |  |
| Female <25, 365 days | Female <25, 365 days | 15.0 | 0.767 (0.730 - 0.805) | 0.450 (0.377 - 0.527) | 3.0 (2.5 -3.6) |
| Full-cohort, 365 days |  | 14.8 | 0.735 (0.692 -0.778) | 0.409 (0.334 -0.482) | 2.8 (2.3 -3.3) |
| Female [25 ,64], 365 days | Female [25 ,64], 365 days | 8.4 | 0.808 (0.784 - 0.832) | 0.409 (0.358 - 0.458) | 4.9 (4.3 -5.5) |
| Full-cohort, 365 days |  | 8.8 | 0.836 (0.815 -0.856) | 0.432 (0.38 -0.485) | 4.9 (4.3 -5.5) |
| Female >64, 365 days | Female >64, 365 days | 3.1 | 0.754 (0.673 - 0.835) | 0.111 (0.054 - 0.236) | 3.6 (2.0 -7.9) |
| Full-cohort, 365 days |  | 3.1 | 0.756 (0.665 -0.848) | 0.115 (0.061 -0.202) | 3.8 (2.3 -6.5) |
| Male <25, 365 days | Male <25, 365 days | 4.6 | 0.737 (0.653 - 0.821) | 0.249 (0.141 - 0.400) | 5.4 (3.3 -8.9) |
| Full-cohort, 365 days |  | 4 | 0.748 (0.666 -0.83) | 0.252 (0.124 -0.394) | 6.3 (3.5 -10.5) |
| Male [25 ,64], 365 days | Male [25 ,64], 365 days | 5.6 | 0.835 (0.801 - 0.859) | 0.444 (0.390 - 0.498) | 8.0 (7.0 -9.2) |
| Full-cohort, 365 days |  | 5.4 | 0.806 (0.779 -0.832) | 0.32 (0.265 -0.373) | 5.9 (4.9 -7.0) |
| Male >64, 365 days | Male >64, 365 days | 3.3 | 0.525 (0.397 - 0.653) | 0.054 (0.021 - 0.177) | 1.6 (0.8 -6.1) |
| Full-cohort, 365 days |  | 3.5 | 0.769 (0.662 -0.876) | 0.227 (0.075 -0.407) | 6.5 (2.5 -13.1) |

Model performance metrics have been evaluated in the test dataset. AUCROC: area under the receiver operating characteristic curve. AUCPR: area under the precision-recall curve. Adjusted AUCPR= AUCPR divided by the prevalence of positive cases.

Across sex, the pooled model achieved similar performance compared with sex-specific models. Specifically, the female-specific model achieved an AUCROC of 0.801 and an adjusted AUCPR of 4.5. The full-cohort model evaluated in female obtained AUCROC of 0.824 and an adjusted AUCPR 4.6. In males, the sex-specific model reached an AUCROC of 0.773 and adjusted AUCPR of 5.4. The full-cohort model in males achieved an AUCROC 0.797and an adjusted AUCPR of 6.0.

In age and sex subgroups, performance patterns differed by sex. Among females, the full-cohort model and the stratified models performed similarly across all age groups. For males, the full-cohort model provided notable improvements in older age group (>64 years), where the stratified model achieved an AUCROC of 0.525 compared with 0.769 in the full-cohort model, and adjusted AUCPR of 1.6 compared with 6.5. In the younger age group in males, the full-cohort model also performed slightly better, with adjusted AUCPR of 6.3 versus 5.4 for the stratified approach. In contrast, in the 25-64 male age group, the stratified approach performed slightly better, with adjusted AUCPR of 8.0 compared with 5.9 for the full cohort model. These results suggest that a 365-day full-cohort model offers robust discrimination and strong relative predictive value across sexes, and age groups, with particular gains in male older adults and shorter prediction windows.

### Model performance across different data splitting strategies

Model performance varied according to the data splitting strategy (**Supplementary Table 7**). The episode-level split achieved the highest overall performance, with an AUCROC of 0.819 (95% CI: 0.805 - 0.834) and an adjusted AUCPR of 5.4 (95% CI: 5.0 -5.9). In contrast, subject-level splitting reduced performance significantly (AUCROC = 0.711, 95% CI: 0.694 -0.729; adjusted AUCPR 2.8, 95% CI: 2.5 -3.1), suggesting that the inclusion of repeated episodes from the same individual in both training and validation sets strongly influences predictive accuracy. A random single-episode-per-subject split yielded a similar performance to subject-level partitioning (AUCROC = 0.754, 95% CI: 0.731 -0.777; adjusted AUCPR = 3.2, 95% CI: 2.7 -3.8). Temporal validation using data from 2015–2017 for training and 2018 for testing resulted in intermediate performance (AUCROC = 0.746, 95% CI: 0.732 -0.76; adjusted AUCPR = 3.1, 95% CI: 2.8 -3.3). Overall, these findings highlight that predictive performance varies substantially depending on the splitting strategy, with episode-level splits producing the most optimistic estimates, while subject-level and temporal splits provide more conservative performance metrics.

## DISCUSSION

Using population-based electronic health record data from Catalonia, Spain, this study developed and evaluated a series of machine learning–based models to predict intentional self-harm following discharge from psychiatric hospitalization. We examined model accuracy and the relative importance of predictors across different follow-up horizons (7 to 365 days), sex, and age groups, assessed the generalizability of a 365-day model to shorter horizons and specific subgroups, and explored how alternative data-splitting strategies affected model robustness and performance.

The episode-level prediction models developed in our study achieved good discrimination, with a median AUCROC of 0.775 (IQR 0.764 to 0.808) across event horizon, sex, and sex-age stratified analyses. Precision recall performance also suggested clinical relevance: the adjusted AUCPR showed a median 5.4-fold improvement over baseline prevalence (IQR 4.5 to 6.2). Notably, the 365-day model trained on the full cohort performed on par with models trained specifically for shorter horizons and for subgroups defined by sex or age. In addition, in strata with low model performance due to low sample size, such as hospitalized males aged 65 years or older in our study, the 365-day overall model substantially improved discrimination (AUCROC 0.77 vs 0.53). This suggests a pragmatic pathway toward a single deployable model for all subgroups, especially when data are sparse. Adoption of such a unified approach should, however, be contingent on calibration and fairness evaluation specific to each subgroup, as undertaken here. While a unified model can perform well overall, subgroup-specific models may better identify the most relevant predictors for each group, which can be useful for guiding individualized clinical decisions. When data are limited, however, a unified model remains a valid and practical option.

Building on these findings, it is important to note that model performance in our study varied markedly by data splitting technique applied in the model validation phase. Random episode level splits (often the standard technique applied in previous studies [69]) yielded the highest apparent performance (AUCROC 0.82); however, this technique breaks temporal order as it allows for models to be trained on future hospitalizations, including future hospitalizations from the same individual, leading to overly optimistic model performance estimates. When we used more conservative strategies (i.e., subject level splitting, selecting a single random episode per person, and temporal splitting), AUCROC declined to the range 0.71 to 0.75, with parallel reductions in AUCPR. Among these, temporal splitting most closely mirrors real clinical deployment within the same health system of which the source data was extracted, where self-harm prediction in a patient is based on historical information from the same patient (including previous hospitalizations) as well as other patients, to predict future self-harm. This approach also implies that the model must be adaptable to potential changes in patient characteristics and clinical practice over time, as these changes may affect its performance [70]. Under temporal validation, performance remained clinically meaningful, with an adjusted AUCPR around 3.1, that is, roughly three times the baseline prevalence. We therefore recommend reporting results under multiple validation schemes and treating episode level estimates as upper bounds, while confirming real world performance through prospective and external evaluation.

When situated in the broader literature, our 365-day episode-level AUCROC of 0.82 aligns closely with the low-bias benchmark (≈0.82) reported in a systematic review of machine learning-based prediction models for suicide-related outcomes though somewhat below the overall pooled estimate of 0.86, which likely reflects optimism from less rigorous model development strategies in previous studies [20]. In comparison with another systematic review [16], which found that many studies in psychiatric populations reported accuracies above 70% but often without external validation or precision-recall-based evaluation, our study adds strength by explicitly incorporating conservative validation and reporting AUCPR adjusted for baseline prevalence. Relative to the only previous benchmark focusing on post-discharge psychiatric hospitalization using Danish registry data [26], which developed separate 30-day models for suicide attempts (AUCROC 0.85) and suicide deaths (AUCROC 0.71), our 30-day model achieved an AUCROC of 0.76, somewhat lower than their model but comparable to their suicide model. At 365 days, our full sample model yielded comparable discrimination (AUCROC 0.82 episode-level split; 0.75 temporal split) and greater generalizability across time horizons and subgroups, consistent with the use of a longer prediction window, composite outcome, and emphasis on prevalence-sensitive metrics. Differences with this Danish study may partly reflect the substantially richer set of sociodemographic and familial predictors available in Danish national registries, including marital status, income, occupational status, parental health history, and education, several of which were among the strongest predictors in their models. Collectively, these comparisons suggest that our estimates are in line with, though somewhat more conservative than, those from prior work and underscore the value of rigorous validation, horizon choice, outcome definition, and predictor availability in shaping model performance.

Our study also provides several important insights into the most important predictor variables for intentional self-harm. First, while previous self-harm was found to be an important predictor in the 90-and 180-day prediction models, none of the self-harm related variables were found to be important in the 365-day models developed, including overall and sex-age stratified models. This is surprising, as previous self-harm is considered among the strongest and most replicated predictors for future self-harm [16,71,72]. One potential explanation could be the use of shorter event horizons (< 6 months) in most prior studies, suggesting that because the strength of autocorrelation between self-harm events decreases over time, prior self-harm events are only relevant in the first weeks and months after discharge, in line with our findings. It has also been shown that the relative importance of predictors varies by horizon: shorter periods are more strongly associated with recent events (e.g., emergency visits, hospitalizations), whereas longer periods rely more on sociodemographic factors and accumulated clinical trajectories [73].

Second, the set of most important predictor variables across models varied greatly by sex-age groups, and to a lesser extent, by event horizon. Especially when focusing on age group-specific models in each sex, very few of the important predictor variables were shared across models. Importantly, our SHAP detailed analyses revealed that the contribution of individual predictors to self-harm risk was not uniform but varied substantially across sex and age strata, and, in some cases, even within strata depending on co-occurring conditions. While certain diagnoses, such as depression, borderline personality disorder, and cocaine and sedative use disorders, adjustment disorders and developmental disorders were almost exclusively associated with elevated risk, in line with previous literature [74–76], other predictors, including schizophrenia-related diagnoses, bipolar disorder, antipsychotic dispensations, mixed anxiety–depression, alcohol and cannabis use disorder, and childhood disorders, demonstrated context-dependent associations, with their influence on risk estimates differing across subgroups and interacting features. These findings highlight the complexity and heterogeneity of risk mechanisms underlying self-harm and illustrate the necessity of modeling high-order interactions to achieve clinically meaningful risk stratification.

Several limitations should be acknowledged when interpreting these findings. First, due to a relatively low number of suicide cases, our outcome combined NLISH and suicide into a single category, which may have introduced heterogeneity given that risk factors for attempts and deaths partly diverge [26,77]. Second, the look-back window for NLISH diagnoses was restricted to one year, imposed by the availability of emergency department and CSRC data only from 2014 onwards. Because previous NLISH is among the strongest predictors of future self-harm, this restriction may have attenuated model performance. Third, our models were restricted to routinely collected registry data. Unlike the Danish national registries used in a previous benchmark study [26] we lacked access to sociodemographic and family history variables such as marital status, income, occupational status, parental health history, and education, which emerged among the most important predictors in this previous study. We also could not incorporate survey-based psychosocial information or unstructured data from clinical notes via natural language processing, limiting the capability to capture risk factors such as acute stressors, hopelessness, or impulsivity. In addition, during the study period (2014–2019), the Catalan health system transitioned from ICD-9-CM to ICD-10-CM, and because our predictor variables were defined through grouped ICD codes, this change may have affected how predictors were constructed and thus affected model inputs and comparability over time, particularly in the temporal split model. Finally, our models were developed and tested within a single regional health system, which may limit generalizability to other settings; thresholds were not calibrated to service capacity; and most importantly, our findings remain retrospective, requiring prospective and external validation before clinical implementation.

Our study demonstrates that machine learning models based on routinely collected health registry data can predict intentional self-harm after psychiatric hospitalization with good discrimination and clinically meaningful precision–recall performance. A unified 365-day model generalized well across horizons and subgroups, offering a pragmatic pathway toward deployable risk stratification, particularly in data-sparse populations. By applying conservative validation strategies, we provide performance estimates that are more likely to reflect real-world use. However, before clinical implementation, external and prospective validation, calibration and fairness assessments, and alignment of thresholds with service capacity will be essential. Future work integrating registry data with socioeconomic information, survey responses, and unstructured clinical notes may further enhance predictive accuracy and clinical utility.

## Data availability

The primary data, including healthcare, mortality, and administrative records, were provided by a third party, the Agency for Quality and Assessment of Catalonia (Agència de Qualitat i Avaluació Sanitàries de Catalunya; AQuAS), under the PADRIS (Programa d’Analítica de Dades per a la Recerca i la Innovació en Salut) framework. Access to these data is restricted, and must comply with PADRIS’s legal and ethical requirements. Interested parties can obtain access to the data, code, and documentation upon request, in accordance with the agreement’s provisions. The minimum dataset needed to replicate the analyses underlying this study, including the anonymized individual-level registry data, data dictionaries, and statistical code, are available upon reasonable request from the corresponding author (Gemma Vilagut), provided (a) the purpose is to replicate our analysis and results without additional investigator support, (b) access is granted following approval of a brief proposal and the signing of a Data Access Agreement, and (c) the request aligns with the terms of our agreement with PADRIS/AQuAS and is approved by the PADRIS legal representative.

## Code availability

The code used for data processing, statistical analyses, and figure generation in this study is available at [GitHub repository link]. The repository contains the full analysis pipeline. Access to the code is unrestricted; however, execution requires the underlying data, which can only be shared under the conditions specified in the Data Availability Statement.

## Acknowledgments

This study was conducted using anonymized data provided by the Agency for Quality and Assessment of Catalonia (Agència de Qualitat i Avaluació Sanitàries de Catalunya; AQuAS), within the framework of the Data Analytics Program for Health Research and Innovation (Programa d’analítica de dades per a la recerca i la innovació en salut; PADRIS) Programme. We would like to thank Juan Francisco Martínez Cerdá, PhD, from the Agency for Health Quality and Assessment of Catalonia (AQuAS), Barcelona, Spain, for his support in providing information on the data sources.

## References

1. Naghavi M. Global, regional, and national burden of suicide mortality 1990 to 2016: systematic analysis for the Global Burden of Disease Study 2016. BMJ 2019 Feb 6;l94. doi: 10.1136/bmj.l94

2. Global burden of 369 diseases and injuries in 204 countries and territories, 1990–2019: a systematic analysis for the Global Burden of Disease Study 2019. Lancet Lond Engl 2020 Oct 17;396(10258):1204–1222. PMID:33069326

3. Fazel S, Runeson B. Suicide. N Engl J Med 2020 Jan 16;382(3):266–274. PMID:31940700

4. An J, Wang Q, Bai Z, Du X, Yu D, Mo X. Global burden and trend of substance use disorders, self-harm, and interpersonal violence from 1990 to 2021, with projection to 2040. BMC Public Health 2025 May 2;25(1):1632. PMID:40317000

5. Xie L, Tang L, Liu Y, Dong Z, Zhang X. Global burden and trends of self-harm from 1990 to 2021, with predictions to 2050. Front Public Health 2025;13:1571579. PMID:40438046

6. Sutar R, Kumar A, Yadav V. Suicide and prevalence of mental disorders: A systematic review and meta-analysis of world data on case-control psychological autopsy studies. Psychiatry Res 2023 Nov;329:115492. PMID:37783094

7. Witt K, Stewart A, Hawton K. Practitioner Review: Treatments for young people who self-harm - challenges and recommendations for research and clinical practice. J Child Psychol Psychiatry 2025 Jan;66(1):122–131. PMID:39194179

8. Witt KG, Hetrick SE, Rajaram G, Hazell P, Salisbury TLT, Townsend E, Hawton K. Psychosocial interventions for self-harm in adults. Cochrane Database Syst Rev John Wiley & Sons, Ltd; 2021;(4). doi: 10.1002/14651858.CD013668.pub2

9. Olfson M, Wall M, Wang S, Crystal S, Liu S-M, Gerhard T, Blanco C. Short-term Suicide Risk After Psychiatric Hospital Discharge. JAMA Psychiatry American Medical Association (AMA); 2016 Nov 1;73(11):1119. doi: 10.1001/jamapsychiatry.2016.2035

10. Dt C, Cj R, D H-P, Sp S, C S, Mm L. Suicide Rates After Discharge From Psychiatric Facilities: A Systematic Review and Meta-analysis. JAMA Psychiatry JAMA Psychiatry; 2017 Jan 7;74(7). PMID:28564699

11. Premature Death, Suicide, and Nonlethal Intentional Self-Harm After Psychiatric Discharge | Psychiatry and Behavioral Health | JAMA Network Open | JAMA Network. Available from: https://jamanetwork.com/journals/jamanetworkopen/fullarticle/2820374 [accessed Jul 27, 2025]

12. O’Connell PH, Durns T, Kious BM. Risk of suicide after discharge from inpatient psychiatric care: a systematic review. Int J Psychiatry Clin Pract 2021 Nov;25(4):356–366. PMID:32749183

13. Carter G, Milner A, McGill K, Pirkis J, Kapur N, Spittal MJ. Predicting suicidal behaviours using clinical instruments: systematic review and meta-analysis of positive predictive values for risk scales. Br J Psychiatry J Ment Sci 2017 Jun;210(6):387–395. PMID:28302700

14. Woodford R, Spittal MJ, Milner A, McGill K, Kapur N, Pirkis J, Mitchell A, Carter G. Accuracy of Clinician Predictions of Future Self-Harm: A Systematic Review and Meta-Analysis of Predictive Studies. Suicide Life Threat Behav 2019 Feb;49(1):23–40. PMID:28972271

15. Bentley KH, Kennedy CJ, Khadse PN, Brooks Stephens JR, Madsen EM, Flics MJ, Lee H, Smoller JW, Burke TA. Clinician Suicide Risk Assessment for Prediction of Suicide Attempt in a Large Health Care System. JAMA Psychiatry 2025 Jun 1;82(6):599–608. doi: 10.1001/jamapsychiatry.2025.0325

16. Pigoni A, Delvecchio G, Turtulici N, Madonna D, Pietrini P, Cecchetti L, Brambilla P. Machine learning and the prediction of suicide in psychiatric populations: a systematic review. Transl Psychiatry Nature Publishing Group; 2024 Mar 9;14(1):140. doi: 10.1038/s41398-024-02852-9

17. Barak-Corren Y, Castro VM, Javitt S, Hoffnagle AG, Dai Y, Perlis RH, Nock MK, Smoller JW, Reis BY. Predicting Suicidal Behavior From Longitudinal Electronic Health Records. Am J Psychiatry American Psychiatric Association Publishing; 2017 Feb 1;174(2):154–162. doi: 10.1176/appi.ajp.2016.16010077

18. Oliver D, Arribas M, Perry BI, Whiting D, Blackman G, Krakowski K, Seyedsalehi A, Osimo EF, Griffiths SL, Stahl D, Cipriani A, Fazel S, Fusar-Poli P, McGuire P. Using Electronic Health Records to Facilitate Precision Psychiatry. Biol Psychiatry Elsevier; 2024 Oct 1;96(7):532–542. PMID:38408535

19. Graham S, Depp C, Lee EE, Nebeker C, Tu X, Kim H-C, Jeste DV. Artificial Intelligence for Mental Health and Mental Illnesses: an Overview. Curr Psychiatry Rep 2019 Nov 7;21(11):116. doi: 10.1007/s11920-019-1094-0

20. Kusuma K, Larsen M, Quiroz JC, Gillies M, Burnett A, Qian J, Torok M. The performance of machine learning models in predicting suicidal ideation, attempts, and deaths: A meta-analysis and systematic review. J Psychiatr Res 2022 Nov 1;155:579–588. doi: 10.1016/j.jpsychires.2022.09.050

21. Marcus SC, Cullen SW, Schmutte T, Xie M, Liu T, Ungar LH, Cardamone NC, Williams NJ, Olfson M. A cohort study of predictors of short-term nonfatal suicidal and self-harm events among individuals with mental health disorders treated in the emergency department. J Psychiatr Res 2025 Jun;186:458–468. PMID:40318538

22. Bayramli I, Castro V, Barak-Corren Y, Madsen EM, Nock MK, Smoller JW, Reis BY. Temporally informed random forests for suicide risk prediction. J Am Med Inform Assoc JAMIA 2021 Nov 2;29(1):62–71. PMID:34725687

23. Wolock CJ, Williamson BD, Shortreed SM, Simon GE, Coleman KJ, Yeargans R, Ahmedani BK, Daida Y, Lynch FL, Rossom RC, Ziebell RA, Cruz M, Wellman RD, Coley RY. Importance of variables from different time frames for predicting self-harm using health system data. medRxiv 2024 Sep 20;2024.04.29.24306260. PMID:39371167

24. Kessler RC, Bauer MS, Bishop TM, Demler OV, Dobscha SK, Gildea SM, Goulet JL, Karras E, Kreyenbuhl J, Landes SJ, Liu H, Luedtke AR, Mair P, McAuliffe WHB, Nock M, Petukhova M, Pigeon WR, Sampson NA, Smoller JW, Weinstock LM, Bossarte RM. Using Administrative Data to Predict Suicide After Psychiatric Hospitalization in the Veterans Health Administration System. Front Psychiatry 2020;11:390. PMID:32435212

25. Mitra A, Chen K, Liu W, Kessler RC, Yu H. Post-discharge suicide prediction among US veterans using natural language processing-enriched social and behavioral determinants of health. Npj Ment Health Res Nature Publishing Group; 2025 Feb 22;4(1):8. doi: 10.1038/s44184-025-00120-2

26. Nielsen SD, Christensen RHB, Madsen T, Karstoft K-I, Clemmensen L, Benros ME. Prediction models of suicide and non-fatal suicide attempt after discharge from a psychiatric inpatient stay: A machine learning approach on nationwide Danish registers. Acta Psychiatr Scand 2023;148(6):525–537. doi: 10.1111/acps.13629

27. Jiang T, Rosellini AJ, Horváth-Puhó E, Shiner B, Street AE, Lash TL, Sørensen HT, Gradus JL. Using machine learning to predict suicide in the 30 days after discharge from psychiatric hospital in Denmark. Br J Psychiatry Cambridge University Press; 2021 Aug;219(2):440–447. doi: 10.1192/bjp.2021.19

28. Gradus JL, Rosellini AJ, Horváth-Puhó E, Jiang T, Street AE, Galatzer-Levy I, Lash TL, Sørensen HT. Predicting Sex-Specific Nonfatal Suicide Attempt Risk Using Machine Learning and Data From Danish National Registries. Am J Epidemiol 2021 Dec 1;190(12):2517–2527. doi: 10.1093/aje/kwab112

29. Tsui F, Ruiz VM, Ryan ND, Shi L, Melhem NM, Young JF, Davis M, Gibbons R, Brent DA. Risk for Suicide Attempts Assessed Using the Patient Health Questionnaire–9 Modified for Teens. JAMA Netw Open 2024 Oct 8;7(10):e2438144. doi: 10.1001/jamanetworkopen.2024.38144

30. McQuaid RJ, Nikolitch K, Vandeloo KL, Burhunduli P, Phillips JL. Sex Differences in Determinants of Suicide Risk Preceding Psychiatric Admission: An Electronic Medical Record Study. Front Psychiatry 2022 May 27;13:892225. PMID:35711595

31. Miranda-Mendizabal A, Castellví P, Alayo I, Vilagut G, Blasco MJ, Torrent A, Ballester L, Almenara J, Lagares C, Roca M, Sesé A, Piqueras JA, Soto-Sanz V, Rodríguez-Marín J, Echeburúa E, Gabilondo A, Cebrià AI, Bruffaerts R, Auerbach RP, Mortier P, Kessler RC, Alonso J. Gender commonalities and differences in risk and protective factors of suicidal thoughts and behaviors: A cross-sectional study of Spanish university students. Depress Anxiety 2019 Nov;36(11):1102–1114. doi: 10.1002/da.22960

32. Miret M, Caballero FF, Huerta-Ramírez R, Moneta MV, Olaya B, Chatterji S, Haro JM, Ayuso-Mateos JL. Factors associated with suicidal ideation and attempts in Spain for different age groups. Prevalence before and after the onset of the economic crisis. J Affect Disord 2014 Jul 1;163:1–9. doi: 10.1016/j.jad.2014.03.045

33. Liu Q, Ren Y, Huang Y, Xie R, Li Y, Dong Z, Zhang X. Development and validation of a probability-assessment nomogram for non-suicidal self-injury in hospitalized adolescents and young adults with mental disorders. Sci Rep Nature Publishing Group; 2025 Apr 30;15(1):15142. doi: 10.1038/s41598-025-00142-y

34. Boudreaux ED, Rundensteiner E, Liu F, Wang B, Larkin C, Agu E, Ghosh S, Semeter J, Simon G, Davis-Martin RE. Applying Machine Learning Approaches to Suicide Prediction Using Healthcare Data: Overview and Future Directions. Front Psychiatry Frontiers Media S.A.; 2021 Aug;12. doi: 10.3389/fpsyt.2021.707916

35. Wang J, Qiu J, Zhu T, Zeng Y, Yang H, Shang Y, Yin J, Sun Y, Qu Y, Valdimarsdóttir UA, Song H. Prediction of Suicidal Behaviors in the Middle-aged Population: Machine Learning Analyses of UK Biobank. JMIR Public Health Surveill 2023 Feb 20;9(1):e43419. doi: 10.2196/43419

36. Wilkinson PO, Qiu T, Jesmont C, Neufeld SAS, Kaur SP, Jones PB, Goodyer IM. Age and gender effects on non-suicidal self-injury, and their interplay with psychological distress. J Affect Disord 2022 Jun 1;306:240–245. doi: 10.1016/j.jad.2022.03.021

37. Zang C, Hou Y, Lyu D, Jin J, Sacco S, Chen K, Aseltine R, Wang F. Accuracy and transportability of machine learning models for adolescent suicide prediction with longitudinal clinical records. Transl Psychiatry Nature Publishing Group; 2024 Jul 31;14(1):316. doi: 10.1038/s41398-024-03034-3

38. Morita K, Mizuno T, Kusuhara H. Investigation of a Data Split Strategy Involving the Time Axis in Adverse Event Prediction Using Machine Learning. J Chem Inf Model 2022 Sep 12;62(17):3982–3992. doi: 10.1021/acs.jcim.2c00765

39. Cho H, She J, De Marchi D, El-Zaatari H, Barnes EL, Kahkoska AR, Kosorok MR, Virkud AV. Machine Learning and Health Science Research: Tutorial. J Med Internet Res 2024 Jan 30;26:e50890. doi: 10.2196/50890

40. Lyu Y, Li H, Sayagh M, Jiang ZM (Jack), Hassan AE. An Empirical Study of the Impact of Data Splitting Decisions on the Performance of AIOps Solutions. ACM Trans Softw Eng Methodol 2021 Jul 23;30(4):54:1-54:38. doi: 10.1145/3447876

41. Programa d’Analítica de Dades per a la Recerca i la Innovació en Salut. Agència Qual Avaluació Sanitàries Catalunya AQuAS. Available from: http://aquas.gencat.cat/ca/fem/intelligencia-analitica/padris/ [accessed Jul 25, 2025]

42. INE. Instituto Nacional de Estadística. INE. Available from: https://www.ine.es/ [accessed Jul 25, 2025]

43. International Classification of Diseases (ICD). Available from: https://www.who.int/standards/classifications/classification-of-diseases [accessed Jul 25, 2025]

44. Allebeck P. The use of population based registers in psychiatric research. Acta Psychiatr Scand Wiley; 2009 Nov;120(5):386–391. doi: 10.1111/j.1600-0447.2009.01474.x

45. ICD - ICD-9-CM - International Classification of Diseases, Ninth Revision, Clinical Modification. 2024. Available from: https://archive.cdc.gov/www_cdc_gov/nchs/icd/icd9cm.htm [accessed Aug 30, 2025]

46. CDC. ICD-10-CM Files. Classif Dis Funct Disabil. 2025. Available from: https://www.cdc.gov/nchs/icd/icd-10-cm/files.html [accessed Aug 30, 2025]

47. CDC. ICD-10. Classif Dis Funct Disabil. 2024. Available from: https://www.cdc.gov/nchs/icd/icd-10/index.html [accessed Sep 3, 2025]

48. Blecker S, Adhikari S, Zhang H, Dodson JA, Desai SM, Anzisi L, Pazand L, Schoenthaler AM, Mann DM. Validation of EHR medication fill data obtained through electronic linkage with pharmacies. J Manag Care Spec Pharm 2021 Oct;27(10):1482–1487. PMID:34595945

49. Anatomical Therapeutic Chemical (ATC) Classification. Available from: https://www.who.int/tools/atc-ddd-toolkit/atc-classification [accessed Jul 25, 2025]

50. Mortier P, Vilagut G, Puértolas Gracia B, De Inés Trujillo A, Alayo Bueno I, Ballester Coma L, Blasco Cubedo MJ, Cardoner N, Colls C, Elices M, Garcia-Altes A, Gené Badia M, Gómez Sánchez J, Martín Sánchez M, Morros R, Prat Pubill B, Qin P, Mehlum L, Kessler RC, Palao D, Pérez Sola V, Alonso J, CODIRISC Epidemiology Study Group. Catalonia Suicide Risk Code Epidemiology (CSRC-Epi) study: protocol for a population-representative nested case-control study of suicide attempts in Catalonia, Spain. BMJ Open 2020 Jul 12;10(7):e037365. PMID:32660952

51. StratifiedKFold. Scikit-Learn. Available from: https://scikit-learn/stable/modules/generated/sklearn.model_selection.StratifiedKFold.html [accessed Aug 15, 2025]

52. Wang C, Wu Q, Weimer M, Zhu E. FLAML: A Fast and Lightweight AutoML Library.

53. Truong A, Walters A, Goodsitt J, Hines K, Bruss CB, Farivar R. Towards Automated Machine Learning: Evaluation and Comparison of AutoML Approaches and Tools. 2019 IEEE 31st Int Conf Tools Artif Intell ICTAI 2019. p. 1471–1479. doi: 10.1109/ICTAI.2019.00209

54. Metin A, Bilgin TT. Automated machine learning for fabric quality prediction: a comparative analysis. PeerJ Comput Sci 2024 Jul 23;10:e2188. PMID:39145237

55. Ke G, Meng Q, Finley T, Wang T, Chen W, Ma W, Ye Q, Liu T-Y. LightGBM: A Highly Efficient Gradient Boosting Decision Tree.

56. Chen T, Guestrin C. XGBoost: A Scalable Tree Boosting System. Proc 22nd ACM SIGKDD Int Conf Knowl Discov Data Min 2016. p. 785–794. doi: 10.1145/2939672.2939785

57. Breiman L. Random Forests. Mach Learn 2001 Oct 1;45(1):5–32. doi: 10.1023/A:1010933404324

58. Geurts P, Ernst D, Wehenkel L. Extremely randomized trees. Mach Learn 2006 Apr 1;63(1):3–42. doi: 10.1007/s10994-006-6226-1

59. Kramer O. K-Nearest Neighbors. In: Kramer O, editor. Dimens Reduct Unsupervised Nearest Neighbors Berlin, Heidelberg: Springer; 2013. p. 13–23. doi: 10.1007/978-3-642-38652-7_2ISBN:978-3-642-38652-7

60. Qin J, Lou Y. L1–2 Regularized Logistic Regression. 2019 53rd Asilomar Conf Signals Syst Comput 2019. p. 779–783. doi: 10.1109/IEEECONF44664.2019.9048830

61. Fawcett T. An introduction to ROC analysis. Pattern Recognit Lett 2006 Jun 1;27(8):861–874. doi: 10.1016/j.patrec.2005.10.010

62. Martin B, Bennett TD, DeWitt PE, Russell S, Sanchez-Pinto LN. Use of the Area Under the Precision-Recall Curve to Evaluate Prediction Models of Rare Critical Illness Events. Pediatr Crit Care Med 2025 Jun;26(6):e855–e859. PMID:40304543

63. Parodi S, Verda D, Bagnasco F, Muselli M. The clinical meaning of the area under a receiver operating characteristic curve for the evaluation of the performance of disease markers. Epidemiol Health Korean Society of Epidemiology; 2022 Oct 17;44:e2022088. doi: 10.4178/epih.e2022088

64. Lundberg S, Lee S-I. A Unified Approach to Interpreting Model Predictions. arXiv; 2017. doi: 10.48550/arXiv.1705.07874

65. Coley RY, Walker RL, Cruz M, Simon GE, Shortreed SM. Clinical risk prediction models and informative cluster size: Assessing the performance of a suicide risk prediction algorithm. Biom J 2021;63(7):1375–1388. doi: 10.1002/bimj.202000199

66. Xue Y, Liang H, Norbury J, Gillis R, Killingworth B. Predicting the risk of acute care readmissions among rehabilitation inpatients: A machine learning approach. J Biomed Inform 2018 Oct 1;86:143–148. doi: 10.1016/j.jbi.2018.09.009

67. Sheu Y, Sun J, Lee H, Castro VM, Barak-Corren Y, Song E, Madsen E, Gordon WJ, Kohane IS, Churchill SE, Reis BY, Cai T, Smoller JW. An Efficient Landmark Model for Prediction of Suicide Attempts in Multiple Clinical Settings. Psychiatry Res 2023 May;323:115175. PMID:37003169

68. Boulitsakis Logothetis S, Green D, Holland M, Al Moubayed N. Predicting acute clinical deterioration with interpretable machine learning to support emergency care decision making. Sci Rep Nature Publishing Group; 2023 Aug 21;13(1):13563. doi: 10.1038/s41598-023-40661-0

69. Lin Y-W, Zhou Y, Faghri F, Shaw MJ, Campbell RH. Analysis and prediction of unplanned intensive care unit readmission using recurrent neural networks with long short-term memory. PLoS ONE 2019 Jul 8;14(7):e0218942. PMID:31283759

70. Vela D, Sharp A, Zhang R, Nguyen T, Hoang A, Pianykh OS. Temporal quality degradation in AI models. Sci Rep Nature Publishing Group; 2022 Jul 8;12(1):11654. doi: 10.1038/s41598-022-15245-z

71. Franklin JC, Ribeiro JD, Fox KR, Bentley KH, Kleiman EM, Huang X, Musacchio KM, Jaroszewski AC, Chang BP, Nock MK. Risk factors for suicidal thoughts and behaviors: A meta-analysis of 50 years of research. Psychol Bull 2017;143(2):187–232. doi: 10.1037/bul0000084

72. Ribeiro JD, Franklin JC, Fox KR, Bentley KH, Kleiman EM, Chang BP, Nock MK. Self-injurious thoughts and behaviors as risk factors for future suicide ideation, attempts, and death: a meta-analysis of longitudinal studies. Psychol Med 2016 Jan;46(2):225–236. doi: 10.1017/S0033291715001804

73. Ahmedani BK, Westphal J, Autio K, Elsiss F, Peterson EL, Beck A, Waitzfelder BE, Rossom RC, Owen-Smith AA, Lynch F, Lu CY, Frank C, Prabhakar D, Braciszewski JM, Miller-Matero LR, Yeh H-H, Hu Y, Doshi R, Waring SC, Simon GE. Variation in Patterns of Health Care Before Suicide: A Population Case-Control Study. Prev Med 2019 Oct;127:105796. PMID:31400374

74. Perea-González MI, De la Vega D, Sanz-Gómez S, Giner L. Personality Disorders and Suicide. A Systematic Review of Psychological Autopsy Studies. Curr Psychiatry Rep 2025 Jan 1;27(1):10–30. doi: 10.1007/s11920-024-01572-7

75. Athey A, Shaff J, Kahn G, Brodie K, Ryan TC, Sawyer H, DeVinney A, Nestadt PS, Wilcox HC. Association of substance use with suicide mortality: An updated systematic review and meta-analysis. Drug Alcohol Depend Rep 2024 Dec 13;14:100310. PMID:39830682

76. Arnone D, Karmegam SR, Östlundh L, Alkhyeli F, Alhammadi L, Alhammadi S, Alkhoori A, Selvaraj S. Risk of suicidal behavior in patients with major depression and bipolar disorder – A systematic review and meta-analysis of registry-based studies. Neurosci Biobehav Rev 2024 Apr 1;159:105594. doi: 10.1016/j.neubiorev.2024.105594

77. Kim E, Han DH, Hwang H, Kim NY, Chung SA, Han L, Kim SM. Comparison of Clinical Indicators for Non-Suicidal Self-Injury and Suicide Attempts in the Emergency Department. J Korean Med Sci 2025 Jul 2;40(33). doi: 10.3346/jkms.2025.40.e205

