## Supplementary materials for "Predicting Intentional Self-Harm Following Psychiatric Discharge in Catalonia, Spain: Machine Learning Models from Linked Registry Data"

**SUPPLEMENTARY MATERIAL**

**Supplementary Table 1. Categories of Mental Disorders, Physical Disorders, and Psychotropic Medications as Predictors of Intentional Self-Harm, with Corresponding ICD-9-CM/ICD-10-CM Diagnostic and ATC Codes.**

| MENTAL AND PHYSICAL DISORDER CATEGORIES | | | |
|---|---|---|---|
| | **ICD9CM** | **ICD10** | **ICD10CM** |
| Acute stress reaction | 308* | F430* | F430* |
| Adjustment disorders | 309, 3090*-3094*, 3098, 30982*-30989*, 3099* | F43, F432*-F439* | F43, F432*-F439* |
| Behavioural and emotional disorders with onset usually occurring in childhood and adolescence | 307,3070*,3072*,3073*,3076*-3077*,3079*,312,3120*-3122*,3124*-3129*,313, 3130*-3133*,3138,31381*-31389*,3139*,314* | F90*, F91*, F92, F928*-F929*, F93*-F95*,F98* | F90*, F91*, F93*-F95*,F98*, Z62891* |
| Bipolar or manic affective disorder | 2960*-2961*, 2964*-2967*, 2968, 29680*, 29681*, 29689* | F30*,F31* | F30*,F31* |
| Borderline personality disorder | 30183* | F603* | F603* |
| Depressive episodes | 2962*, 2980*, 29682*, 311* | F32* | F32, F320*-F325*, F328, F3289*, F329* |
| Diseases of the circulatory system (selection) | 401, 4010*, 4011*-4019*, 402*-405*, 410*, 412*-414*,415, 4150*, 4151, 41512*-41519*, 416*-417*, 420*-429*,430*-438*, 441*,447, 4470*-4476*, 4478*-4479*, V1255* | I10*,I11*-I15*,I20*, I21*-I22*,I23*-I25*,I26*-I28*,I30*-I51*,I60*-I69*,I71*, I77* | I10*, I11*-I16*, I674*,I20*,I21*-I22*,I23*-I25*, I26*-I28*, I30*-I51*,I60*-I69*, I71*, I77, I770*-I776*, I778, I7789*, I779*, N262*, Z86711* |

| | | | |
|---|---|---|---|
| Diseases of the digestive system (selection) | 07020*-07023*, 0703*, 07032*-07033*, 07044*, 07054*, 4560*-4562*, 571*, 5723*, 5728*, 555*-556*,5641*,571, 5710*-5714*, 57140*-57141*, 57149*, 5715*-5716*, V427* | B18*, I85*, K50*-K51*, K58*, K70*, K72*-K74*, K76*, K754*, Z944* | B18*, I85*, K50*-K51*,K58*, K70*, K72*-K74*, K754*, K76*, Z944* |
| Diseases of the musculoskeletal system and connective tissue | 710*-739* | M00*-M03*, M05*-M99* | M00*-M19*, M1A*-M25*, M30*-M99* |
| Diseases of the respiratory system (selection) | 490*-492*, 494*, 496*,493* | J40*-J47* | J40*-J45*, J47* |
| Disorders of psychological development | 299*, 315, 3150*-3159* | F80*,-F84*, F88*,F89* | F80*-F84*,F88*,F89* |
| Disorders of sexual preference | 3021*-3024*, 30281*-30284*, 30289*, 3029* | F641*, F65* | F641*, F65* |
| Dissociative (conversion) disorders | 2982*, 3001* | F44* | F44* |
| Eating disorders | 3075, 30750*, 30752*-30759*,3071*,30751* | F50, F500*-F509* | F50, F500*, F502*, F508*-F509* |
| Endocrine, nutritional and metabolic diseases (selection) | 249*-250*,260*, 263*,276, 2760*-2765*, 2766, 27669*, 2767*-2769*,278,2780*,9951* | E10*-E14*,E40*, E44*-E46*,E66*,E86*-E87* | E08*-E13*,E40*, E44*-E46*,E66*,E86*, E87, E870*-E876*, E877, E8770*, E8779*, E878* |
| Epilepsy | 345* | G40*-G41* | G40* |
| External causes of morbidity (selection) | E800*-E848*,E880*-E888*,E910*, E9290*-E9291*,E960*-E969* | V01*-V99*, Y85*,W00*-W19*,W65*-W74*,X85*-X99*, Y00*-Y09*, Y871* | V01*-V99*,W00*-W19*,W65*-W74*,X92*-X99*, Y00*-Y09* |

| | | | |
|---|---|---|---|
| Gender identity disorders | 3025*-3026*, 30285* | F64, F640*, F642*-F649* | F64, F640*, F642*-F649* |
| Generalized anxiety disorder | 30002* | F411* | F411* |
| Habit and impulse disorders | 3123* | F63* | F63* |
| Malignant neoplasms | 140*-149*,150*,151*,152*,153*-154*,155*, 156*, 157*, 158*-171*, 173*-176*, 179*-182*,1830*, 1832*-1839*, 184*-188*, 1890*, 1891*-1899*, 190*-201*, 202, 2020*-2022*, 2023*-2029*, 203*-208*, V1007* | C00*-C34*, C37*-C41*, C44*-C86*,C88*-C90*, C96*, C97*, C91*-C95* | C00*-C34*, C37*-C41*, C44*-C4A*, C50*,C86*,C88*-C90*-C96*, Z8505*, |
| Mental and behavioural disorders associated with the puerperium not elsewhere classified | 6484* | F53* | F53* |
| Mental and behavioural disorders due to use of alcohol | 291, 2911*-2913*, 2915*, 2918, 29182*-29189*, 2919* | F10, F105*-F109* | F10, F109* |
| Mental and behavioural disorders due to use of other, unspecified, or | 292, 2921*, 2928, 29282*-29289*, 2929* | F11, F115*-F119*, F12, F125*-F129*, F13, F135*-F139*, F14, F145*-F149*, F15, F155*-F159*, F16, F165*-F169*, F18, F185*-F189*, F19, F195*-F199* | F11, F119*, F12, F129*, F13, F139*, F14, F149*, F15, F159*, F16, F169*, F18, F189*, F19, F199* |

| | | | |
|---|---|---|---|
| multiple drugs or psychoactive substances | | | |
| Mental disorder, not otherwise specified | 3009* | F99* | F99* |
| Mental retardation | 317*-319* | F70*-F79* | F70*-F79* |
| Mixed anxiety and depressive disorders, inc. other specified and unspecified types | 3000, 30009 | F41, F412-F419 | F41, F413-F419 |
| Multiple sclerosis | 340* | G35* | G35* |
| Nonorganic sleep disorders | 3074* | F51* | F51* |
| Obsessive-compulsive disorder | 3003* | F42* | F42* |
| Organic, including symptomatic, mental disorders | 0461*, 2791*, 290, 2900*-2904*, 2908*-2909*, 293, 2930*-2931*, 2938, 2939*, 29381*-29382*, 29383*-29389*, 294, 2940*, 2941*, 2942*, 2948*-2949*, 310, 3100*-3102*, 3108*-3109*, 332, 3310*, 3311*, 3320*, 3334* | A810*, B220*, F02*,F00*, F01*,F03*-F07*, F09*,G10*, G20*,G30*,G310*, | A810*, F02*,F01*,F03*,F04*,F05*,F06*, F09*, F482*,F07*, G10*, G20*, G310*, |
| Other disorders of adult personality and behaviour | V402*-V403*, V611* | F68* | F68* |
| Other mood (affective) | 2969* | F38*-F39* | F39* |

| | | | |
|---|---|---|---|
| disorder + Unspecified mood (affective) disorder | | | |
| Other nonorganic psychotic disorders | 298, 2981* | F28* | F28* |
| Other personality disorders | 301, 3011*, 30122*, 3019*,3010*,3012, 30120*-30121*, 3013*-3017*, 3018, 30181*, 30182*, 30184*-30189* | F60, F609*, F61*,F600*-F602*,F604*-F608* | F60, F609*,F600*-F602*,F604*-F608* |
| Other schizophrenia spectrum disorders | 297, 2970*- 2973*, 2978*,2979*, 2983*-2988*,30122* | F21*-F24* | F21*-F24* |
| Pain symptoms | 339*, 346, 3460*-3465*, 3467*-3469*, 7840*,7841*,7865, 78650*- 78652*, 78659*,7890*, 7896* | G430*-G439*, G44*,R070*-R074*,R10*, R51*,R52* | G430*-G435*, G437*-G43D*, G44*, G89*, R51*,R070*,R071*, R072*, R078, R0781*-R0789*, R079*,R10, R100*-R103*, R108, R1081*-R1082*, R1084*, R109*, R52* |
| Panic disorder | 30001*, 30021* | F410* | F410* |
| Persistent mood (affective) disorders | 3004*, 3011* | F34*, F920* | F34* |
| Phobic anxiety disorders | D3002,D30020*-30023*,30029* | F40,F400*,F401*,F402*,F408*-F409* | F40,F400*,F401*,F402*, F408*-F409* |
| Post-traumatic stress disorder | 30981* | F431* | F431* |
| Premenstrual dysphoric disorder | 6254* | N943* | F3281*, N943* |

| | | | |
|---|---|---|---|
| Psychological and behavioural disorders associated with sexual development and orientation | 3020* | F66* | F66* |
| Psychological or behavioural factors associated with disorders or diseases classified elsewhere | 316* | F54* | F54* |
| Recurrent depressive disorder | 2963* | F33* | F33* |
| Schizoaffective disorders | 2957* | F25* | F25* |
| Schizophrenia | 295, 2950*-2956*, 2958*-2959* | F20, F200*-F203*, F205*-F209* | F20,F200*-F203*,F205*,F208*-F209* |
| Screening and history of mental health and substance abuse codes | 3051*, 33392*, 7903*, V11*, V154*, V1582*, V6285*, V663*, V701*-V702*, V710*, V79* | R780*-R786*, Z046*, Z134*, Z865*, Z914* | R780*-R786*, Z046*, Z1330*-Z1339*, Z134*, Z7281*, Z865*, Z87891*, Z914* |
| Sexual dysfunction, not caused by organic disorder or disease | 3027*, 30651* | F52* | F52* |

| | | | |
|---|---|---|---|
| Somatoform or neurotic disorders | 3005*-3008*, 306, 3060*-3064*, 3065, 30650*, 30652*-30659*, 3066*-3069*, 3078* | F45*,F48*, F59* | F45*,F48, F481*, F488*-F489*, F59* |
| Substance use disorder - alcohol | 2910*, 2914*,29181*, 303*, 3050* | F100*-F104* | F101*,F102* |
| Substance use disorder - cannabinoids | 3043*, 3052* | F120*-F124* | F121*,F122* |
| Substance use disorder - cocaine | 3042*, 3056* | F140*-F144* | F141*,F142* |
| Substance use disorder - hallucinogens | 3045*,3053* | F160*-F164* | F161*,F162* |
| Substance use disorder - opioids | 3040*,3055* | F110*-F114* | F111*,F112* |
| Substance use disorder - other stimulants, including caffeine | 3044*,3057* | F150*-F154* | F151*,F152* |
| Substance use disorder - other, unspecified, or multiple drugs or psychoactive substances | 2920*, 2922*, 29281*,3046*-3049*, 305, 3058*-3059* | F180*-F184*, F190*-F194*, F55* | F181*, F182*, F191*,F192*, F55* |
| Substance use disorder - sedatives or hypnotics | 3041*,3054* | F130*-F134* | F131*,F132* |

| | | | |
|---|---|---|---|
| Substance use disorder - tobacco | 3051* | F17* | F17* |
| Unspecified disorder of adult personality and behaviour | V409* | F69* | F69* |
| Unspecified nonorganic psychosis | 2989* | F29* | F29* |
| **PSYCHOTROPIC MEDICATION CATEGORIES** | | | |
| | **ATC code** | | |
| Addiction medicine | N05CM02, N07BA03, N07BB01, N07BB02, N07BB03, N07BB04, N07BB05, N07BC01, N07BC02, N07BC51 | | |
| Addiction medicine / SGAs / anti-EPS | N05AL03 | | |
| Analgesics | N02BA01, N02BA11, N02BA51, N02BB02, N02BB51, N02BE01, N02BE51, N02BE71, N02BG07, N02BG08, N02BG10 | | |
| Anti-ADHD | N06BA01, N06BA02, N06BA04, N06BA07, N06BA09, N06BA11, N06BA12 | | |
| Anti-dementia | N06BX03, N06BX08, N06BX13, N06DA02, N06DA03, N06DA04, N06DX01, N06DX02 | | |
| Anti-epileptic | N03AB02, N03AB05, N03AD01, N03AF03, N03AF04, N03AG02, N03AG04, N03AG06, N03AX03, N03AX10, N03AX14, N03AX15, N03AX17, N03AX18, N03AX21, N03AX22, N03AX23 | | |
| Anti-epileptic / anxiolytic | N03AE01 | | |
| Anti-epileptic / mood stabilizer | N03AF01, N03AF02, N03AG01, N03AX09 | | |
| Anti-epileptic / mood stabilizer / addiction medicine | N03AX11 | | |
| Anti-EPS | N04AA01, N04AA02, N04AA04, N04AA05, N04AB02, N07XX06 | | |

| | |
|---|---|
| Anti-migraine | N02CA01, N02CA04, N02CA07, N02CA52, N02CA72, N02CC01, N02CC02, N02CC03, N02CC04, N02CC05, N02CC06, N02CC07, N02CX01, N02CX02 |
| Anti-motion sickness | N05CM05 |
| Anti-parkinson | N04BA02, N04BA03, N04BB01, N04BC01, N04BC02, N04BC04, N04BC05, N04BC06, N04BC07, N04BC09, N04BD01, N04BD02, N04BD03, N04BX01, N04BX02, N04BX04 |
| Anti-vertigo | N07CA01, N07CA02, N07CA03, N07CA52 |
| Anxiolytic | N03AA02, N03AA03, N05BA01, N05BA02, N05BA04, N05BA05, N05BA06, N05BA08, N05BA09, N05BA10, N05BA12, N05BA14, N05BA21, N05BA24, N05BC01, N05BE01, N05CC01, N05CD01, N05CD02, N05CD03, N05CD05, N05CD06, N05CD08, N05CD09, N05CD10, N05CD11, N05CF01, N05CF02, N05CF03, N05CM06 |
| Anxiolytic / anti-motion sickness | N05BB01 |
| Atypical AD | N06AF03, N06AF04, N06AG02, N06AX01, N06AX02, N06AX03, N06AX05, N06AX06, N06AX11, N06AX12, N06AX18, N06AX22, N06AX26 |
| FGAs | N05AA01, N05AA02, N05AA04, N05AA06, N05AB01, N05AB02, N05AB03, N05AB04, N05AB06, N05AC01, N05AC02, N05AC04, N05AD01, N05AD03, N05AD05, N05AD08, N05AF01, N05AF03, N05AF05, N05AG02 |
| Mood stabilizer | N05AN01 |
| Mood stabilizer / addiction medicine / anxiolytic | N02BF01, N02BF02 |
| Mood stabilizer / addiction medicine / anxiolytic | N02BF02 |
| Opioid analgesic | N02AA01, N02AA03, N02AA05, N02AA55, N02AA59, N02AB01, N02AB02, N02AB03, N02AC04, N02AC54, N02AD01, N02AG01, N02AG02, N02AG04, N02AJ06, N02AJ07, N02AJ08, N02AJ09, N02AJ13, N02AJ14, N02AX02, N02AX06 |
| Opioid analgesic / addiction medicine | N02AE01 |

| | |
|---|---|
| Other | N05CH01, N05CM09, N06BC01, N06BX06, N06CA01, N06CA02, N07AA01, N07AA02, N07AA30, N07AA51, N07AB01, N07AB02, N07AX01, N07BA01, N07XX02, N07XX04, N07XX05, N07XX07, N07XX08, N07XX59 |
| SGAs | N05AE03, N05AE04, N05AE05, N05AH01, N05AH02, N05AH03, N05AH04, N05AH05, N05AH06, N05AL01, N05AL05, N05AX08, N05AX12, N05AX13, N05AX15, N05AX16 |
| SNRIs | N06AX16, N06AX17, N06AX21, N06AX23 |
| SSRIs | N06AX24, N06AB03, N06AB04, N06AB05, N06AB06, N06AB08, N06AB10 |
| TCAs | N06AA02, N06AA04, N06AA06, N06AA07, N06AA09, N06AA10, N06AA11, N06AA12, N06AA21, N06AX14 |

**Supplementary Table 2. Number of Psychiatric Hospitalizations per Patient (n=41,827; 2015-2018).**

| Number of psychiatric hospitalizations | n | % |
|---|---|---|
| 1 | 27,712 | 66.2 |
| 2 | 7,758 | 18.5 |
| 3 | 2,993 | 7.2 |
| 4 | 1,465 | 3.5 |
| 5 | 762 | 1.8 |
| 6 | 406 | 1.0 |
| 7 | 222 | 0.5 |
| 8 | 152 | 0.4 |
| 9 | 87 | 0.2 |
| 10 | 70 | 0.2 |
| >=11 | 200 | 0.5 |
| Total | 41,827 | 100.0 |

**Supplementary Table 3. Characteristics of Psychiatric Hospitalizations in the Study Cohort (n=41,827; 2015–2018). Overall and by Sex.**

| | **All hospitalizations n=71,865** | **Hospitalizations among males n=38,032 (52.9%)** | **Hospitalizations among females n=33,833 (47.1%)** | **P-value*** |
|---|---|---|---|---|
| | n (%) | n (%) | n (%) | |
| **Age at discharge** | | | | <0.001 |
| <25 | 11273 (15.7%) | 5540 (14.6%) | 5733 (16.9%) | |
| 25-64 | 51418 (71.5%) | 29117 (76.6%) | 22301 (65.9%) | |
| 65+ | 9174 (12.8%) | 3375 (8.9%) | 5799 (17.1%) | |
| **Current NLISH diagnosis**** | | | | <0.001 |
| No | 69640 (96.9%) | 37201 (97.8%) | 32439 (95.9%) | |
| Yes | 2225 (3.1%) | 831 (2.2%) | 1394 (4.1%) | |
| **Number of previous psychiatric hospitalizations** | | | | <0.001 |
| 0 | 31231 (43.5%) | 16254 (42.7%) | 14977 (44.3%) | |
| 1 | 14450 (20.1%) | 7807 (20.5%) | 6643 (19.6%) | |
| 2 | 8208 (11.4%) | 4472 (11.8%) | 3736 (11.0%) | |
| 3 | 5128 (7.1%) | 2804 (7.4%) | 2324 (6.9%) | |
| 4 | 3330 (4.6%) | 1841 (4.8%) | 1489 (4.4%) | |
| 5 or more | 9518 (13.2%) | 4854 (12.8%) | 4664 (13.8%) | |
| **Number of ED visits for self-harm prior to index psychiatric hospitalization** | | | | <0.001 |
| 0 | 68541 (95.4%) | 36818 (96.8%) | 31723 (93.8%) | |
| 1 | 2747 (3.8%) | 1052 (2.8%) | 1695 (5.01%) | |
| 2 | 390 (0.5%) | 118 (0.3%) | 272 (0.8%) | |
| 3 | 107 (0.2%) | 32 (0.08%) | 75 (0.2%) | |
| 4 | 36 (0.05%) | 10 (0.03%) | 26 (0.08%) | |
| 5 or more | 44 (0.06%) | 2 (0.01%) | 42 (0.1%) | |
| **Intentional self-harm at 365 days follow-up** | | | | <0.001 |
| Yes | 4901 (6.8%) | 1983 (5.2%) | 2918 (8.6%) | |
| No | 66964 (93.2) | 36049 (94.8%) | 30915 (91.4%) | |

*P-value is based on the chi-squared test.

** Current NLISH diagnosis: a binary indicator of whether the diagnosis was recorded for the current (index) psychiatric hospitalization.

**Supplementary Table 4. Outcome distribution by event horizon, overall, and stratified by sample characteristics. Percentage of hospitalizations that are followed by NLISH in each follow up time horizon by sample characteristics groups (*n* = 71,865).**

| | **Proportion of NLISH by time horizon** | | | | |
|---|---|---|---|---|---|
| | **7 days**<br><br>**n=505 (0.7%)** | **30 days**<br><br>**n=1210 (1.7%)** | **90 days**<br><br>**n=2265 (3.1%)** | **180 days**<br><br>**n=3355 (4.7%)** | **365 days**<br><br>**n=4901 (6.8%)** |
| | n (%) | n (%) | n (%) | n (%) | n (%) |
| **Sex** | | | | | |
| Female | 317 (0.9%) | 723 (2.1%) | 1367 (4.0%) | 2044 (6.0%) | 2918 (8.6%) |
| Male | 188 (0.5%) | 487 (1.3%) | 898 (2.4%) | 1311 (3.5%) | 1983 (5.2%) |
| **Age at discharge** | | | | | |
| <25 | 100 (0.9%) | 232 (2.1%) | 493 (4.4%) | 777 (6.9%) | 1116 (9.9%) |
| 25-64 | 342 (0.7%) | 854 (1.7%) | 1602 (3.1%) | 2348 (4.6%) | 3493 (6.8%) |
| 65+ | 63 (0.7%) | 124 (1.3%) | 170 (1.8%) | 230 (2.5%) | 292 (3.2%) |
| **Current NLISH diagnosis*** | | | | | |
| No | 396 (0.6%) | 995 (1.4%) | 1927 (2.8%) | 2913 (4.2%) | 4331 (6.2%) |
| Yes | 109 (4.9%) | 215 (9.7%) | 338 (15.2%) | 442 (19.9%) | 570 (25.6%) |
| **Number of previous psychiatric hospitalizations** | | | | | |
| 0 | 259 (0.8%) | 568 (1.8%) | 932 (3.0%) | 1278 (4.1%) | 1749 (5.6%) |
| 1 | 74 (0.5%) | 203 (1.4%) | 395 (2.7%) | 617 (4.3%) | 930 (6.4%) |
| 2 | 47 (0.6%) | 109 (1.3%) | 237 (2.9%) | 390 (4.7%) | 585 (7.1%) |
| 3 | 29 (0.6%) | 74 (1.4%) | 174 (3.4%) | 266 (5.2%) | 389 (7.6%) |
| 4 | 20 (0.6%) | 56 (1.7%) | 112 (3.4%) | 182 (5.5%) | 281 (8.4%) |
| 5 or more | 76 (0.8%) | 200 (2.1%) | 415 (4.4%) | 622 (6.5%) | 967 (10.2%) |
| **Number of ED visits for self-harm prior to index psychiatric hospitalization** | | | | | |
| 0 | 397 (0.6%) | 941 (1.4%) | 1761 (2.6%) | 2636 (3.9%) | 3935 (5.7%) |
| 1 | 75 (2.7%) | 182 (6.6%) | 347 (12.6%) | 501 (18.2%) | 702 (25.6%) |
| 2 | 17 (4.4%) | 43 (11.0%) | 82 (21.0%) | 113 (29.0%) | 148 (37.9%) |
| 3 | 6 (5.6%) | 14 (13.1%) | 35 (32.7%) | 52 (48.6%) | 57 (53.3%) |
| 4 | 2 (5.6%) | 9 (25.0%) | 12 (33.3%) | 18 (50.0%) | 21 (58.3%) |
| 5 or more | 8 (18.2%) | 21 (47.7%) | 28 (63.6%) | 35 (79.5%) | 38 (86.4%) |

* Current NLISH diagnosis: a binary indicator of whether the diagnosis was recorded for the current (index) psychiatric hospitalization.

**Supplementary Table 5. Parameter settings of best performing models, by data splitting technique, event horizon, and subgroup.**

| Data split | Event horizon | Subgroup | Model | Hyper-parameters |
|---|---|---|---|---|
| Episode | 365 days | Full sample | LightGBM | n_estimators: 902, num_leaves: 48, min_child_samples: 4, learning_rate: 0.030355799620251655, log_max_bin: 5, colsample_bytree: 0.523151241941435, reg_alpha: 0.005791376463241912, reg_lambda: 0.032635531515107036 |
| Episode | 180 days | Full sample | LightGBM | n_estimators: 123, num_leaves: 287, min_child_samples: 42, learning_rate: 0.03517142804002899, log_max_bin: 10, colsample_bytree: 0.7008472550973778, reg_alpha: 0.0010338209185993944, reg_lambda: 0.0009844371893970828 |
| Episode | 90 days | Full sample | Extra Trees | n_estimators: 117, max_features: np.float64(0.08053295714216195), max_leaves: 581, criterion: entropy |
| Episode | 30 days | Full sample | LightGBM | n_estimators: 64, num_leaves: 754, min_child_samples: 115, learning_rate: np.float64(0.05874144356444483), log_max_bin: 10, colsample_bytree: np.float64(0.587474711886233), reg_alpha: np.float64(0.0073263895682540645), reg_lambda: np.float64(0.02237339566679511) |
| Episode | 7 days | Full sample | XGBoost | n_estimators: 128, max_depth: 9, min_child_weight: np.float64(0.01684308247284819), learning_rate: np.float64(0.5713112670120598), subsample: 1.0, colsample_bylevel: 1.0, colsample_bytree: np.float64(0.8574219079068361), reg_alpha: np.float64(0.008893866816114756), reg_lambda: np.float64(70.636434578407) |
| Episode | 365 days | Female | LightGBM | n_estimators: 502, num_leaves: 223, min_child_samples: 8, learning_rate: np.float64(0.009906171996458144), log_max_bin: 10, colsample_bytree: np.float64(0.6605918740036356), reg_alpha: np.float64(0.0012880021429652639), reg_lambda: 0.0009765625 |
| Episode | 365 days | Male | XGBoost | n_estimators: 74, max_depth: 6, min_child_weight: np.float64(1.384993820373257), learning_rate: np.float64(0.23848550326498433), subsample: np.float64(0.7181281802322108), colsample_bylevel: 1.0, colsample_bytree: 1.0, reg_alpha: np.float64(0.01612226133150949), reg_lambda: np.float64(2.460614179855463) |
| Episode | 365 days | Female <25 | LightGBM | n_estimators: 104, num_leaves: 40, min_child_samples: 35, learning_rate: np.float64(0.047612420256211346), log_max_bin: 6, colsample_bytree: np.float64(0.6345005458229007), reg_alpha: np.float64(0.01791517108668702), reg_lambda: np.float64(0.022726014919459765) |
| Episode | 365 days | Female 25-64 | LightGBM | n_estimators: 516, num_leaves: 337, min_child_samples: 5, learning_rate: np.float64(0.009976647114487829), log_max_bin: 10, colsample_bytree: np.float64(0.6300335284257583), reg_alpha: np.float64(0.005655022820750134), reg_lambda: np.float64(0.006312507271793466) |
| Episode | 365 days | Female >65 | LightGBM | n_estimators: 52, num_leaves: 4, min_child_samples: 25, learning_rate: np.float64(0.19175942226378845), log_max_bin: 5, colsample_bytree: |

| | | | | |
|---|---|---|---|---|
| | | | | np.float64(0.6923626203934599), reg_alpha: np.float64(0.3434969196466506), reg_lambda: np.float64(0.10187194944074617) |
| Episode | 365 days | Male <25 | LightGBM | n_estimators: 12, num_leaves: 321, min_child_samples: 5, learning_rate: np.float64(0.48562410728243766), log_max_bin: 6, colsample_bytree: np.float64(0.6015217653040057), reg_alpha: np.float64(0.0019258956051423927), reg_lambda: np.float64(0.08675274518245718) |
| Episode | 365 days | Male 25-64 | Extra Trees | n_estimators: 186, max_features: np.float64(0.06482037235521644), max_leaves: 1714, criterion: np.str_(entropy) |
| Episode | 365 days | Male >65 | XGBoost | n_estimators: 12, max_depth: 5, min_child_weight: np.float64(0.5485418442637724), learning_rate: np.float64(0.19742461130206032), subsample: np.float64(0.7814703476675395), colsample_bylevel: np.float64(0.8380290924664978), colsample_bytree: 1.0, reg_alpha: np.float64(0.15284320948411595), reg_lambda: np.float64(1.5227686668586295) |
| By episode train 2015-2017, test 2018 | 365 days | Full sample | LightGBM | n_estimators: 228, num_leaves: 118, min_child_samples: 7, learning_rate: np.float64(0.0679639318187909), log_max_bin: 9, colsample_bytree: np.float64(0.7920493145586404), reg_alpha: np.float64(1.144015643397468), reg_lambda: np.float64(0.10065013497162789) |
| By subject | 365 days | Full sample | LightGBM | n_estimators: 2969, num_leaves: 125, min_child_samples: 2, learning_rate: np.float64(0.03054537243913895), log_max_bin: 9, colsample_bytree: np.float64(0.5735193704249335), reg_alpha: np.float64(0.08920153750911039), reg_lambda: np.float64(0.010423092295135727) |
| Random episode per subject | 365 days | Full sample | XGBoost | n_estimators: 940, max_leaves: 4, min_child_weight: np.float64(0.08134238888152051), learning_rate: np.float64(0.0291617479728941), subsample: np.float64(0.7902420373104145), colsample_bylevel: np.float64(0.7287755465237779), colsample_bytree: np.float64(0.6532295318867453), reg_alpha: np.float64(0.1327987330196317), reg_lambda: np.float64(1.7195377924684372) |

LightGBM: Light Gradient Boosting Machine, Extra Trees: Extremely Randomized Trees

**Supplementary Table 6. Shapley additive explanation (SHAP) values for prediction models by sex.**

| | Male | Female |
|---|---|---|
| Schizophrenia -recency | 0.29287 | 0.00365 |
| Depressive episode -recency | 0.12500 | 0.00794 |
| Adjustment disorders -recency | 0.11821 | 0.00826 |
| SSRIs -recency | 0.10795 | 0.00208 |
| Mixed anxiety-depression -recency | 0.10458 | 0.00464 |
| Other personality disorders -recency | 0.11494 | 0.00383 |
| Borderline PD -recency | | 0.00847 |
| SSRIs -yes/no | | 0.00317 |
| Eating disorders -recency | | 0.00302 |
| External causes -yes/no | | 0.00389 |
| Unspecified mental disorder -recency | | 0.00214 |
| Other schizophrenia disorders -recency | | 0.00283 |
| Bipolar disorder -recency | | 0.00385 |
| Recurrent depression -recency | | 0.00242 |
| Depressive episode -number | | 0.00253 |
| Anxiolytics -recency | 0.13054 | |
| External causes -recency | 0.14183 | |
| External causes -number | 0.23164 | |
| Childhood-onset disorders -recency | 0.18013 | |
| MH/SUD screening/history -recency | 0.10245 | |
| Alcohol use disorder -number | 0.13862 | |
| Cannabis use disorder -recency | 0.11194 | |
| Cocaine use disorder -recency | 0.12155 | |
| Musculoskeletal diseases -recency | 0.11829 | |

The 15 most relevant shaps were selected for each model and the table was then sorted by the number of relevant predictors across all models. Each column shows SHAP values for specific outcome; therefore, values are not directly comparable across columns, and a higher value in one column does not imply greater relative risk than in another.

**Supplementary Table 7. Model performance across different data splitting strategies.**

| Split | Outcome Prevalence | AUCROC (95% CI) | AUCPR (95% CI) | Adjusted AUCPR (95% CI) |
|---|---|---|---|---|
| Episode-level | 6.8 | 0.819 (0.805 - 0.834) | 0.370 (0.338 - 0.401) | 5.4 (5.0 -5.9) |
| Subject-level | 6.6 | 0.711 (0.694 -0.729) | 0.187 (0.164 -0.21) | 2.8 (2.5 -3.1) |
| Random episode per subject | 5.3 | 0.754 (0.731 -0.777) | 0.169 (0.141 -0.201) | 3.2 (2.7 -3.8) |
| 2015–2017 vs. 2018 | 7.6 | 0.746 (0.732 -0.76) | 0.233 (0.213 -0.255) | 3.1 (2.8 -3.3) |

AUCROC: area under the receiver operating characteristic curve; AUCPR: area under the precision-recall curve; Adjusted AUCPR= AUCPR divided by the prevalence of positive cases.

Subject-level split, grouping all episodes of the same individual together; (ii) temporal splits, where earlier years were used for training and subsequent years for validation (2015–2017 vs. 2018); and (iii) random selection of a single episode per subject

**Supplementary Figure 1. (A) The receiver operating characteristics curves and the area under the receiver operating characteristic curves (AUROC) for intentional self-harm over event horizons. (B) The precision-recall curves and the area under the precision-recall curves for intentional self-harm over event horizons of the models.**

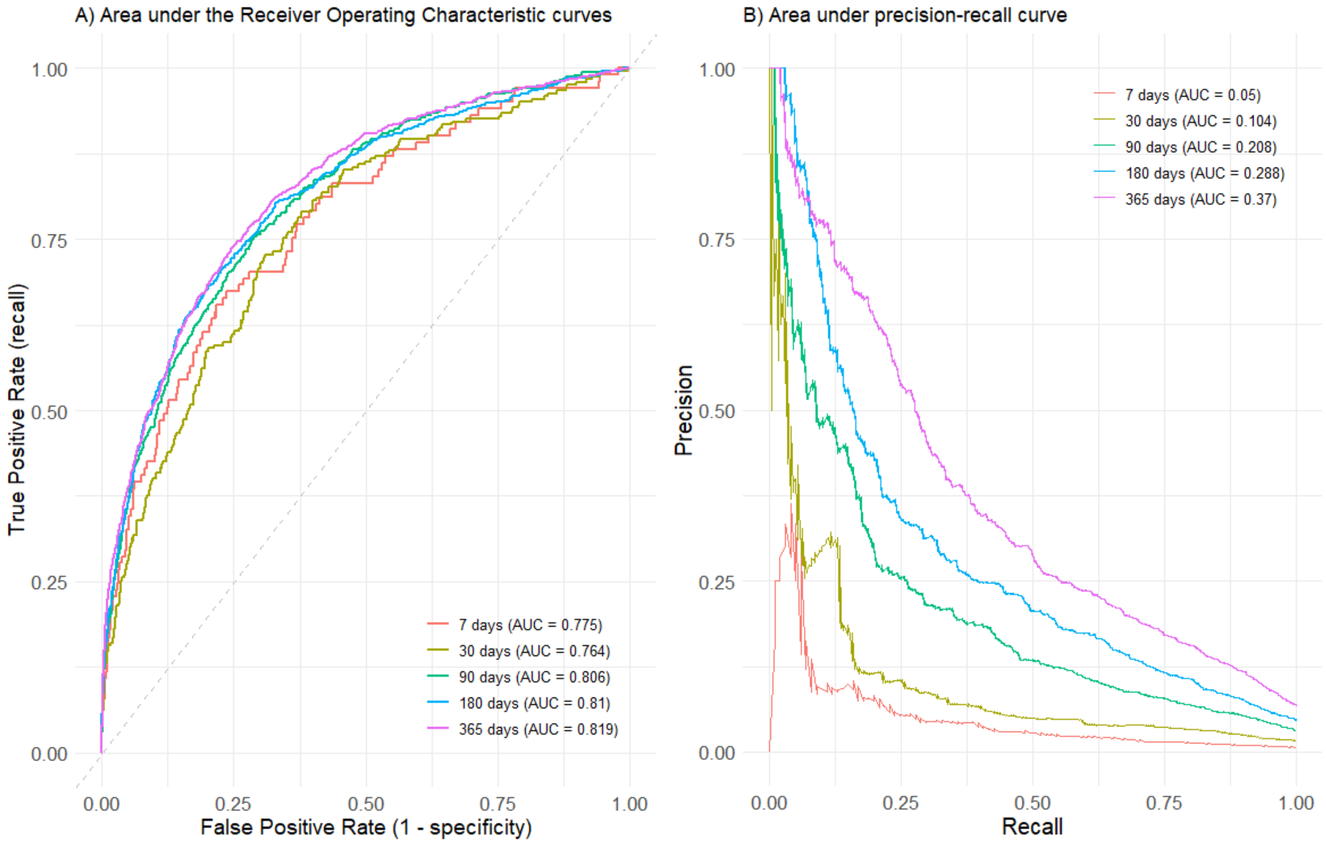

**Supplementary Figure 2. Shapley additive explanation (SHAP) summary graph for prediction models by event horizon. Each point on the graph is a SHAP value for one variable. The colour represents the value of the variable from low (blue) to high (pink).**

**365 days**

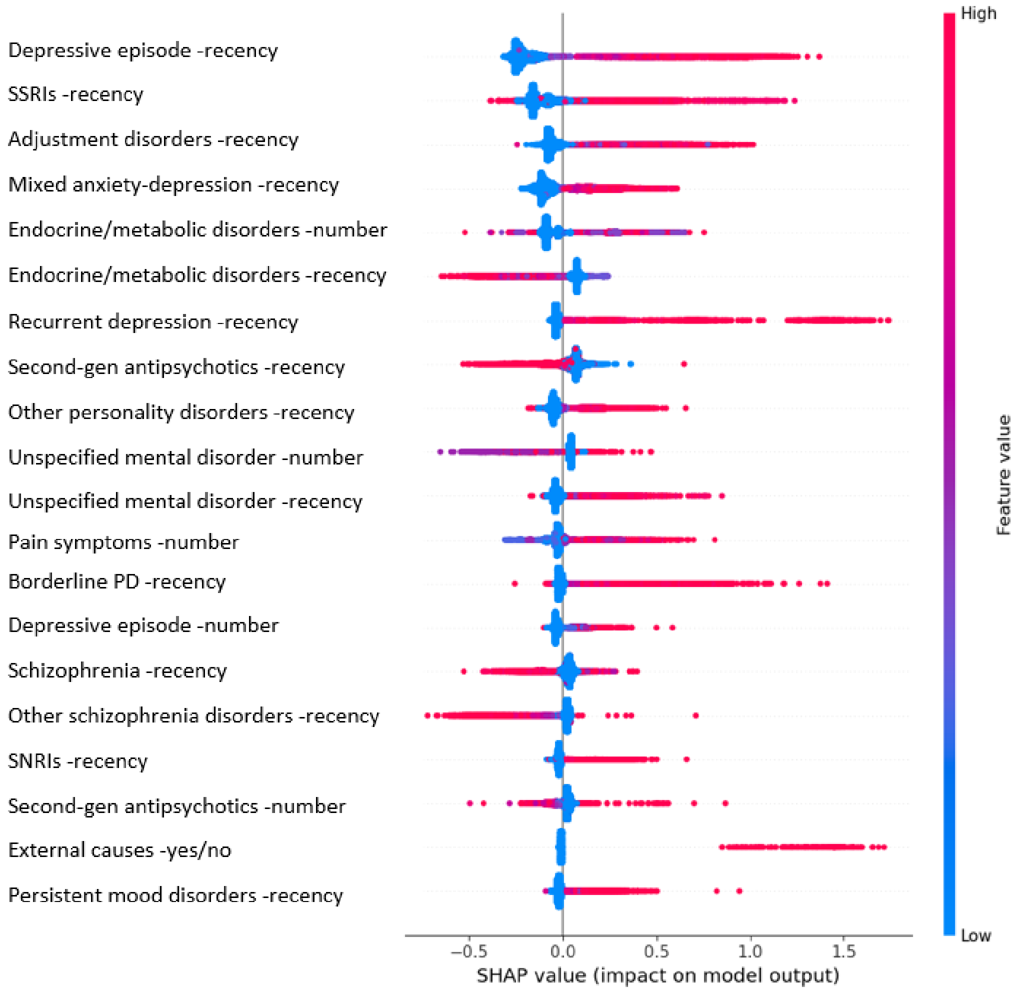


The colour of each point on the graph represents the value of the corresponding variable: pink indicates high values and blue indicates low values. The horizontal axis (x-axis) represents the SHAP value.

**180 days**

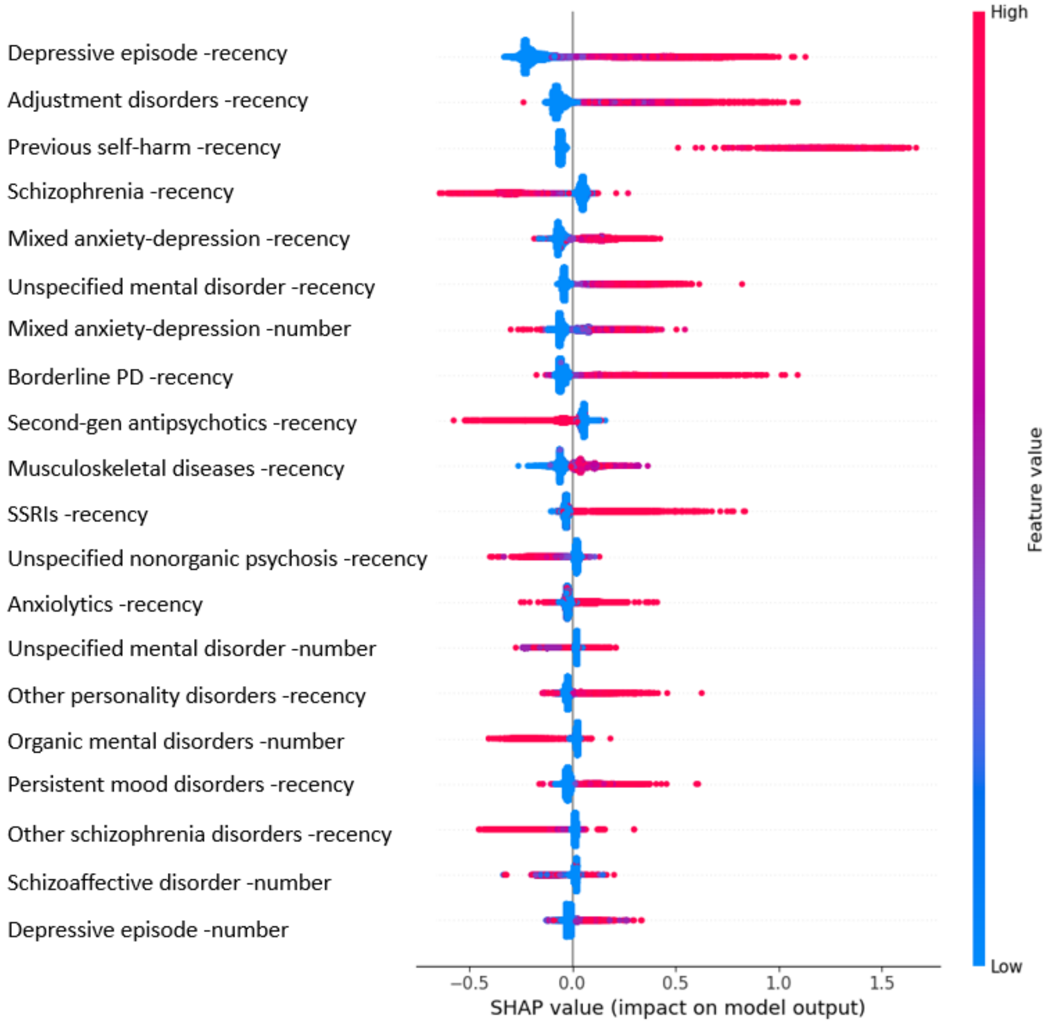


The colour of each point on the graph represents the value of the corresponding variable: pink indicates high values and blue indicates low values. The horizontal axis (x-axis) represents the SHAP value.

**90 days**

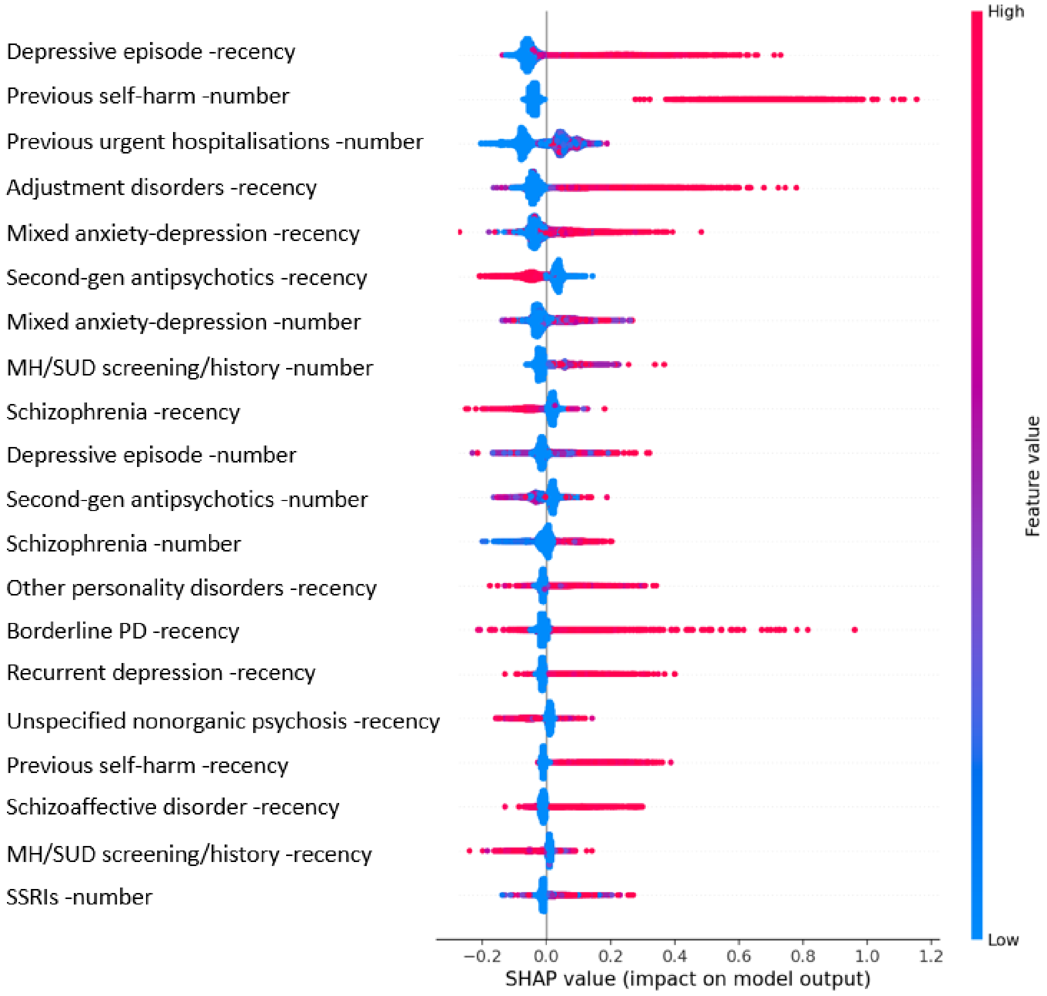


The colour of each point on the graph represents the value of the corresponding variable: pink indicates high values and blue indicates low values. The horizontal axis (x-axis) represents the SHAP value.

**30 days**

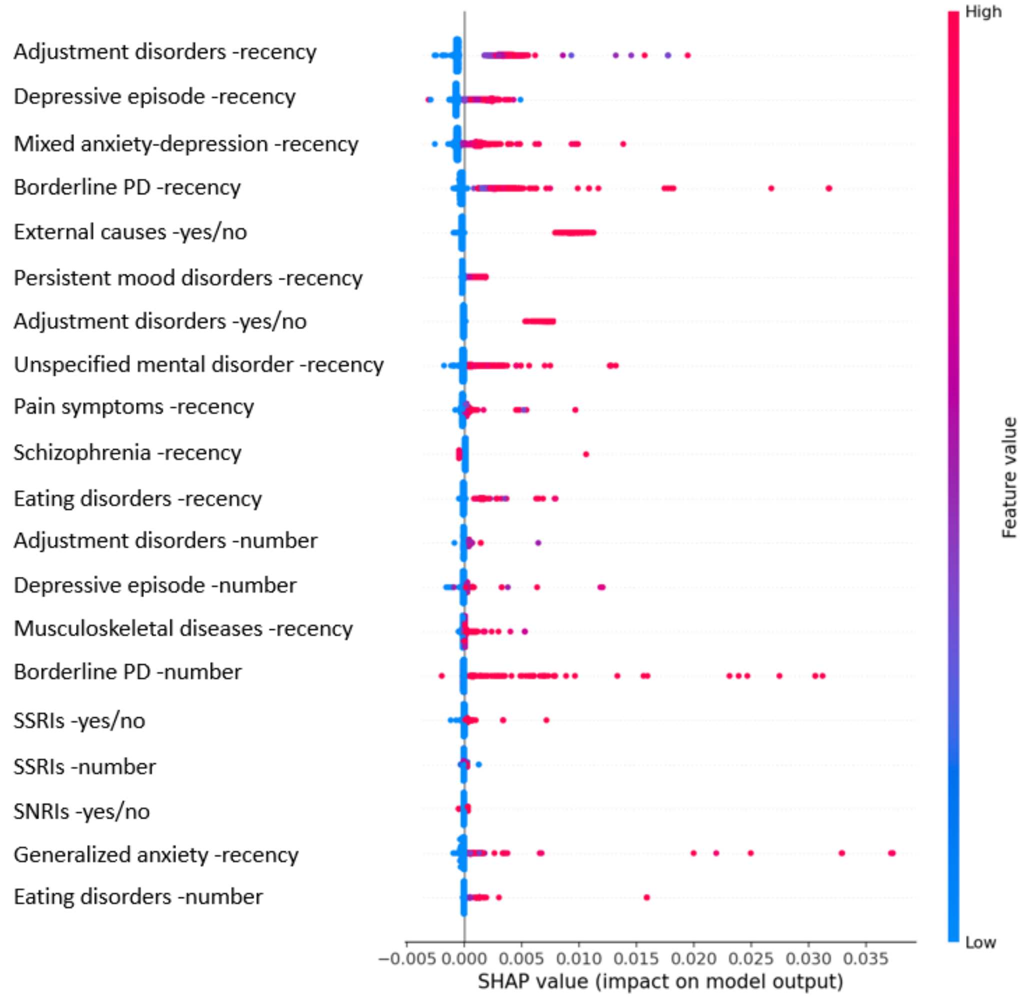


The colour of each point on the graph represents the value of the corresponding variable: pink indicates high values and blue indicates low values. The horizontal axis (x-axis) represents the SHAP value.

**7 days**

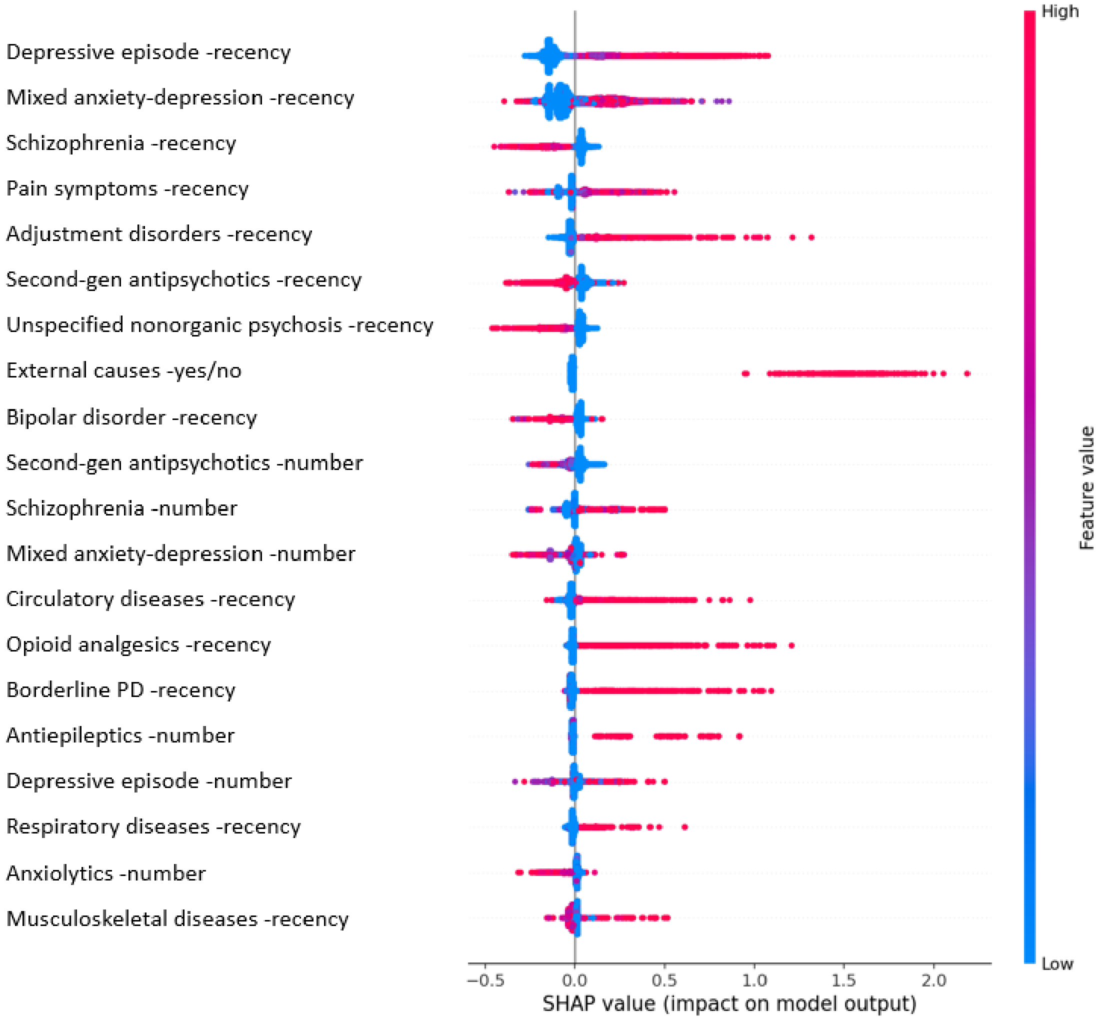


The colour of each point on the graph represents the value of the corresponding variable: pink indicates high values and blue indicates low values. The horizontal axis (x-axis) represents the SHAP value.

**Supplementary Figure 3: Shapley additive explanation (SHAP) summary graph for prediction models by sex. Each point on the graph is a SHAP value for one variable. The colour represents the value of the variable from low (blue) to high (pink).**

**Female**

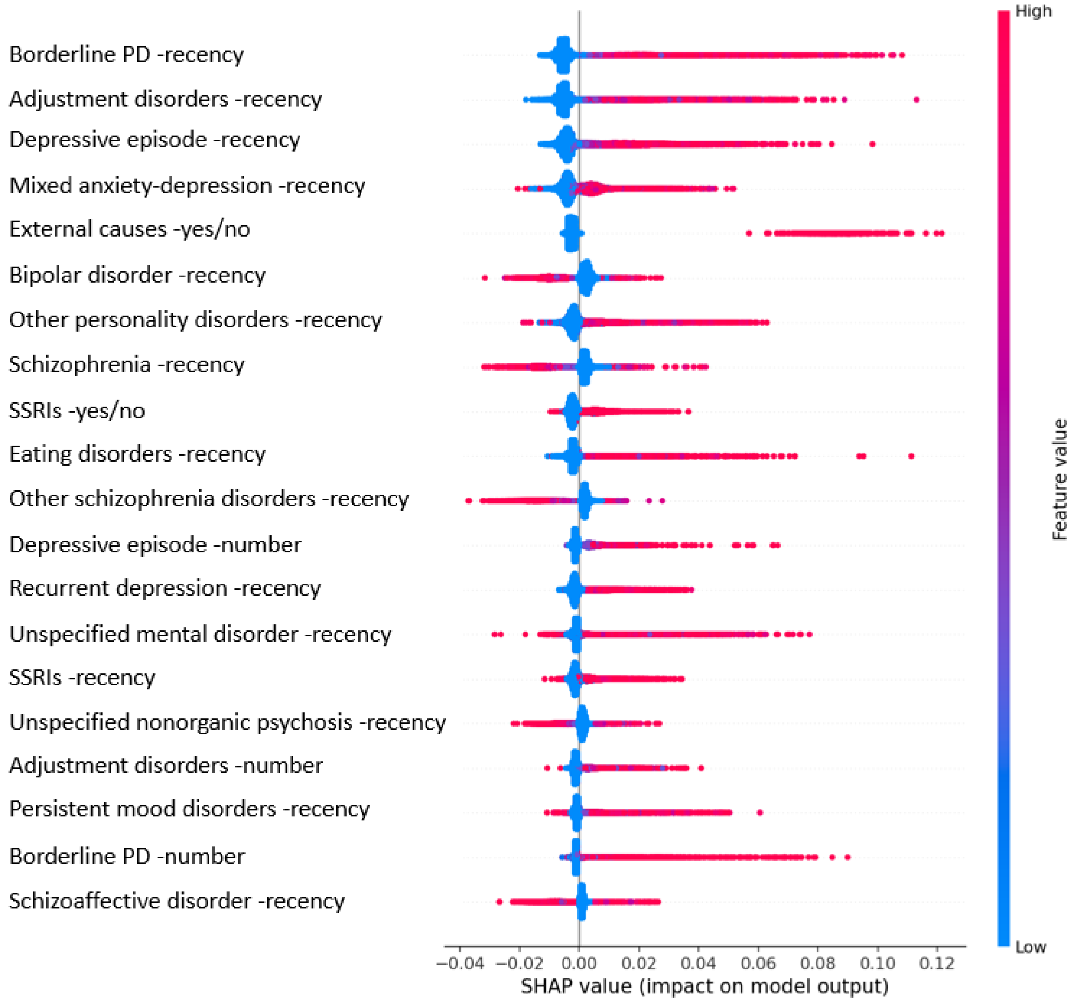


The colour of each point on the graph represents the value of the corresponding variable: pink indicates high values and blue indicates low values. The horizontal axis (x-axis) represents the SHAP value.

**Male**

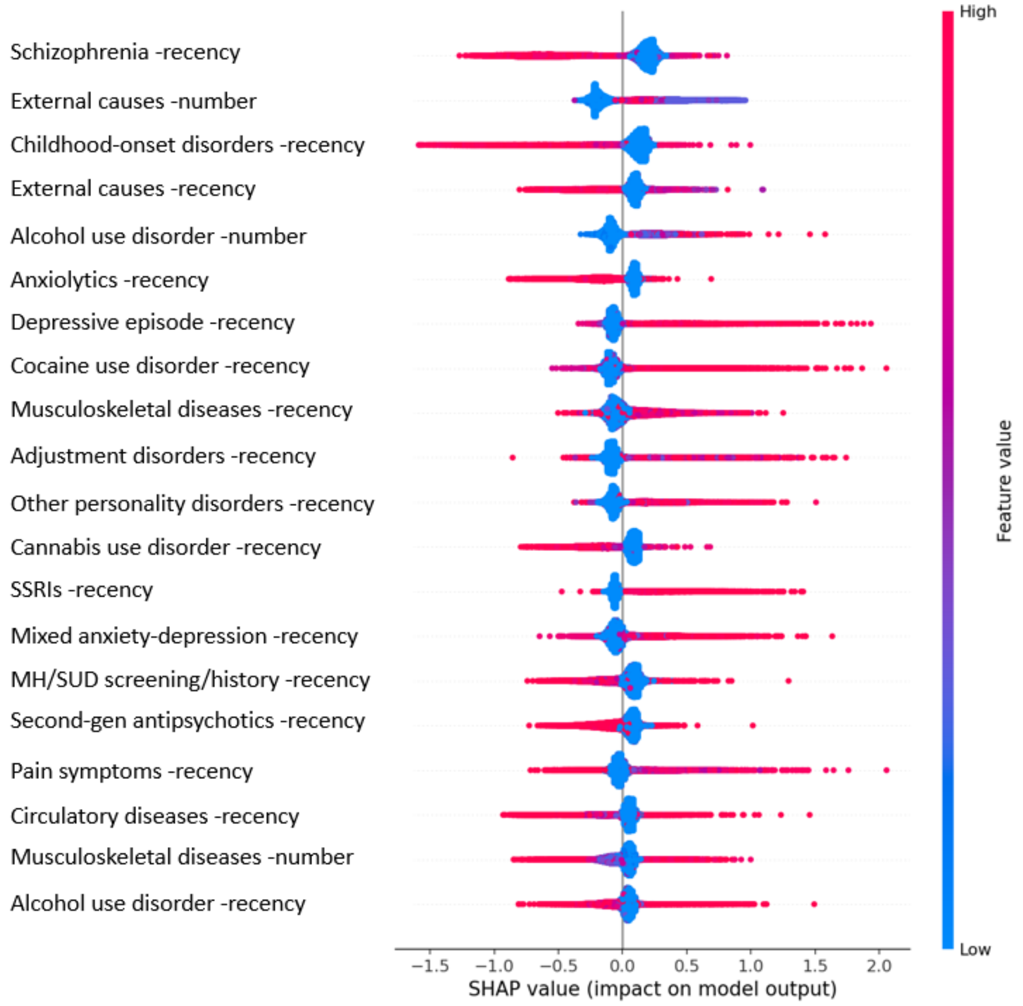


The colour of each point on the graph represents the value of the corresponding variable: pink indicates high values and blue indicates low values. The horizontal axis (x-axis) represents the SHAP value.

**Supplementary Figure 4: Shapley additive explanation (SHAP) summary graph for prediction models by female and age. Each point on the graph is a SHAP value for one variable. The colour represents the value of the variable from low (blue) to high (pink).**

**Female <25y**

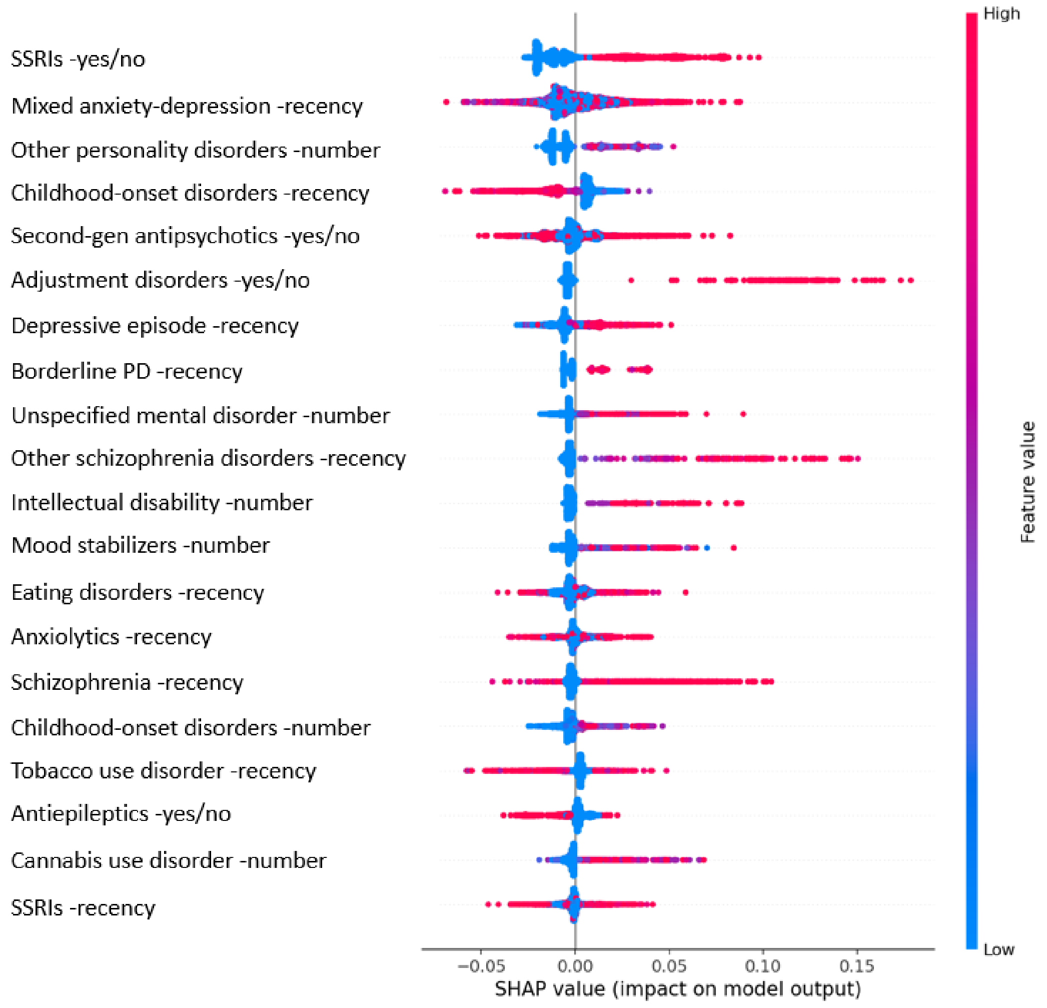


The colour of each point on the graph represents the value of the corresponding variable: pink indicates high values and blue indicates low values. The horizontal axis (x-axis) represents the SHAP value.

**Females 25-64y**

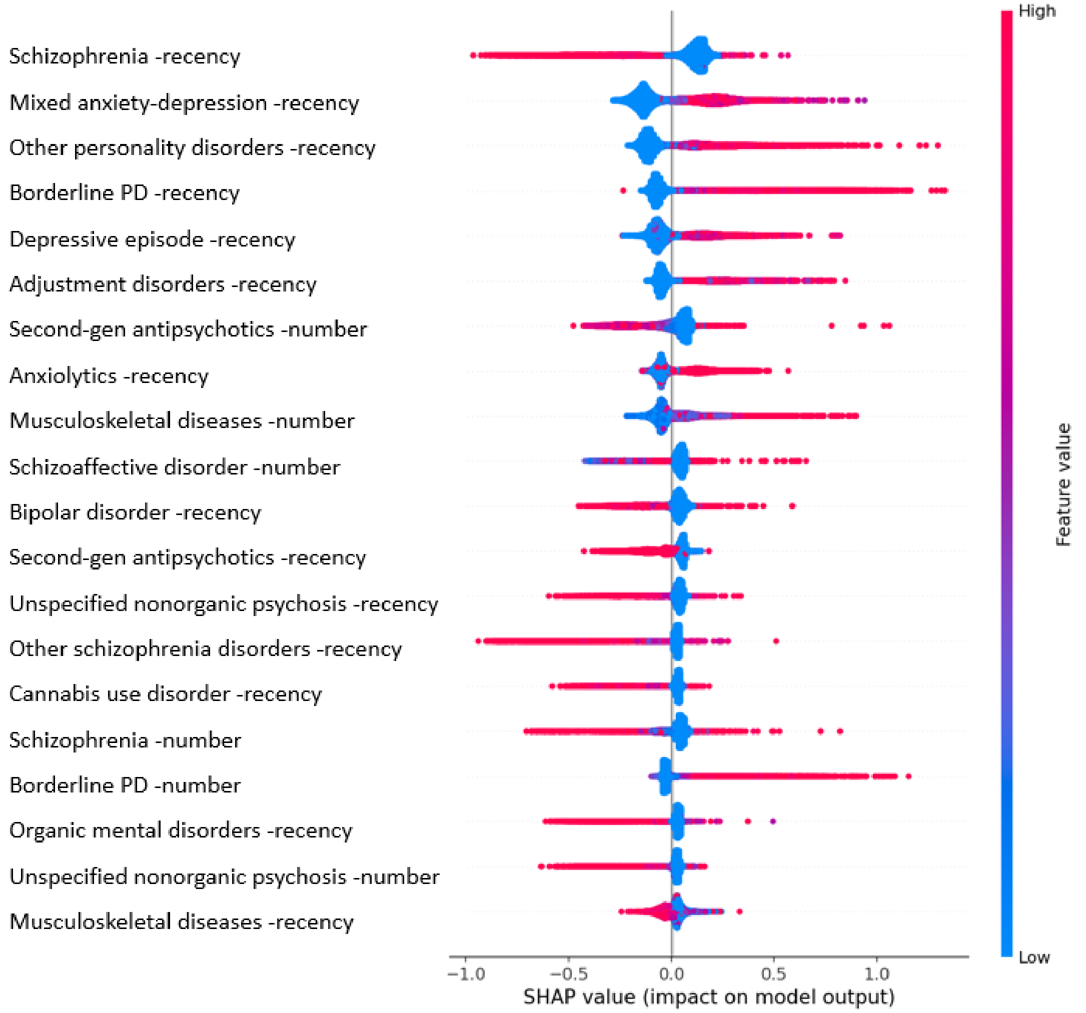


The colour of each point on the graph represents the value of the corresponding variable: pink indicates high values and blue indicates low values. The horizontal axis (x-axis) represents the SHAP value.

**Females 65+y**

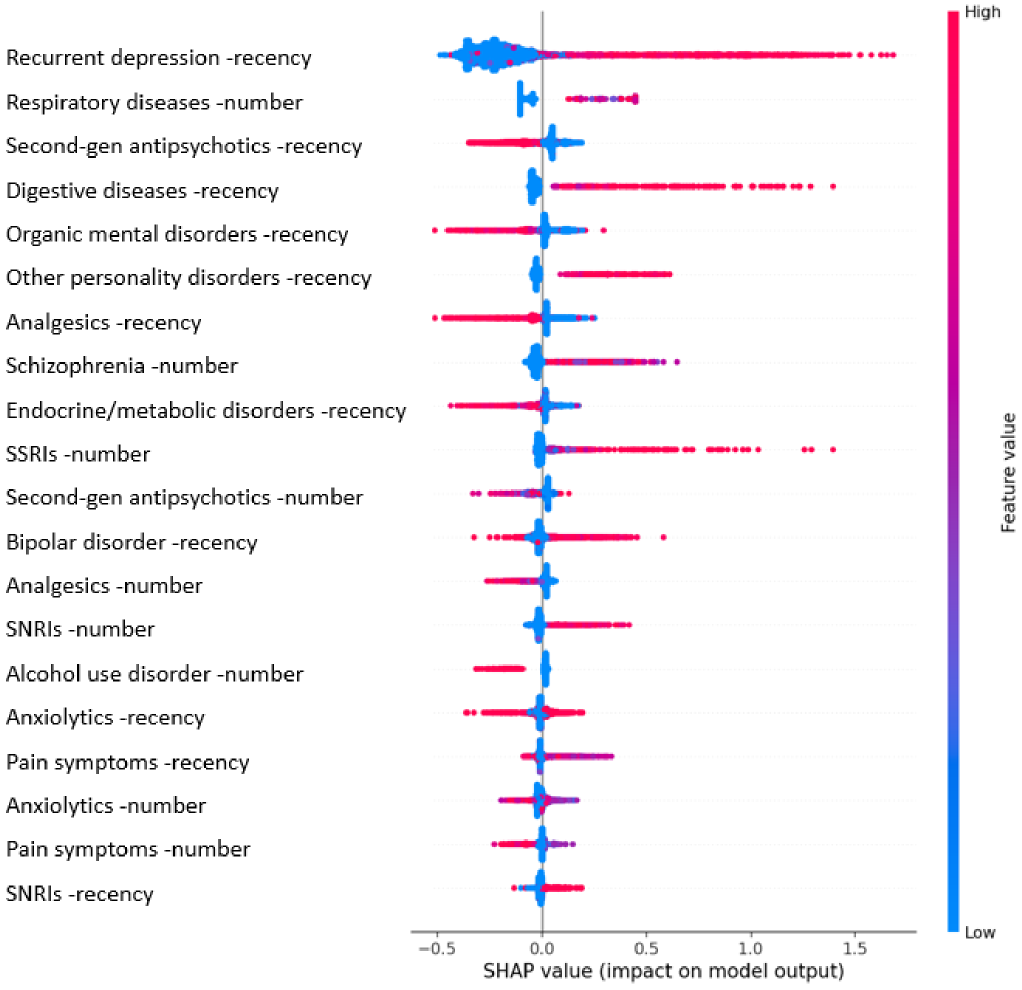


The colour of each point on the graph represents the value of the corresponding variable: pink indicates high values and blue indicates low values. The horizontal axis (x-axis) represents the SHAP value.

Supplementary **Figure 5: Shapley additive explanation (SHAP) summary graph for prediction models by male and age. Each point on the graph is a SHAP value for one variable. The colour represents the value of the variable from low (blue) to high (pink).**

**Male <25y**

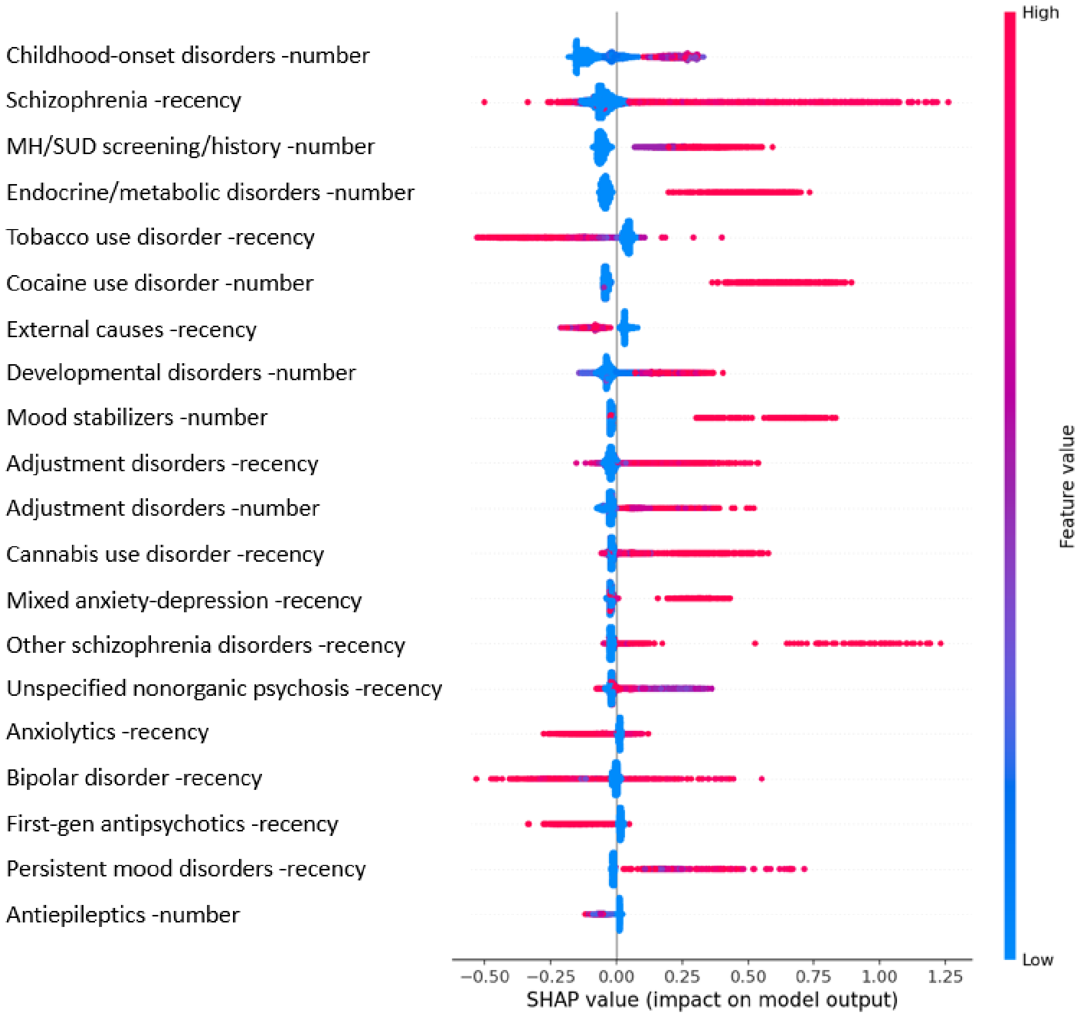


The colour of each point on the graph represents the value of the corresponding variable: pink indicates high values and blue indicates low values. The horizontal axis (x-axis) represents the SHAP value.

**Males 25-64y**

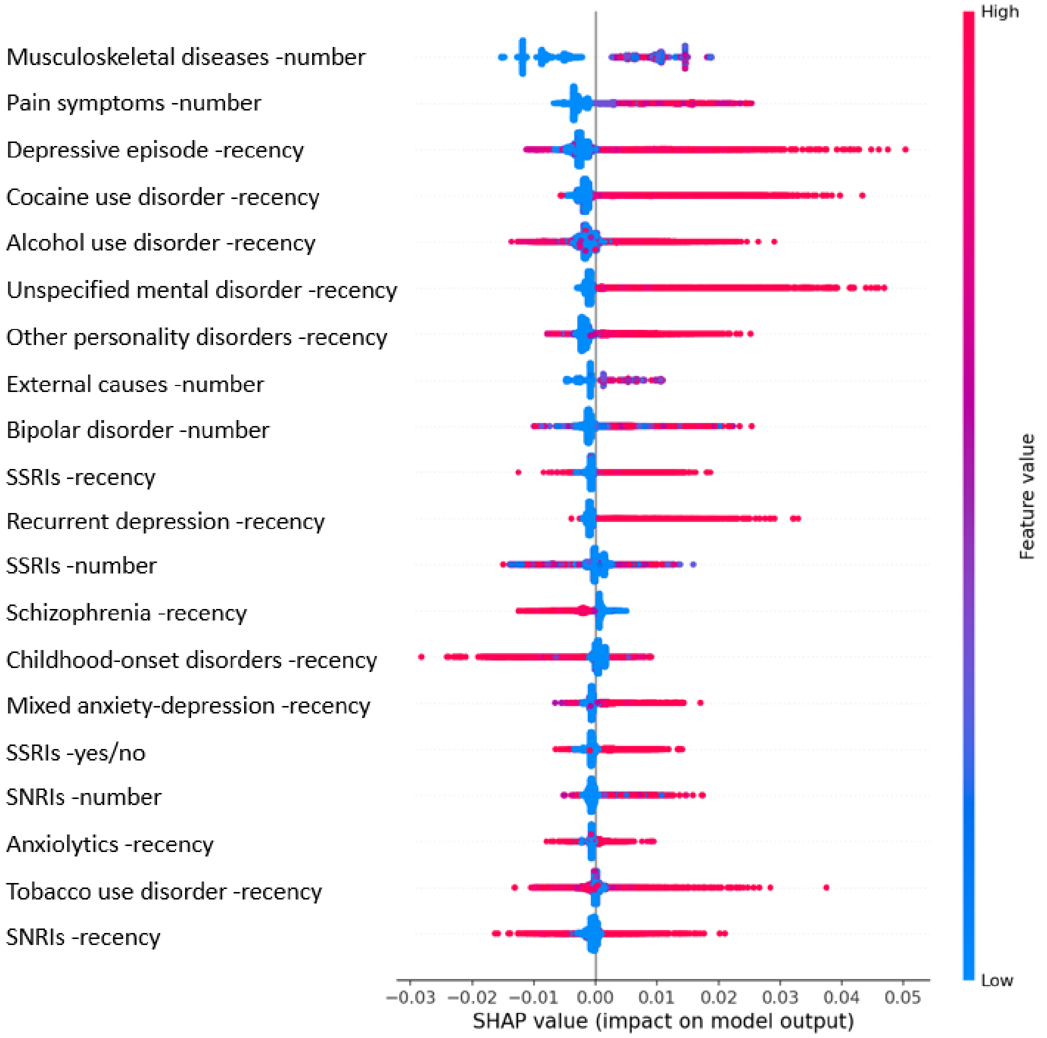


The colour of each point on the graph represents the value of the corresponding variable: pink indicates high values and blue indicates low values. The horizontal axis (x-axis) represents the SHAP value.

**Males 65+y**

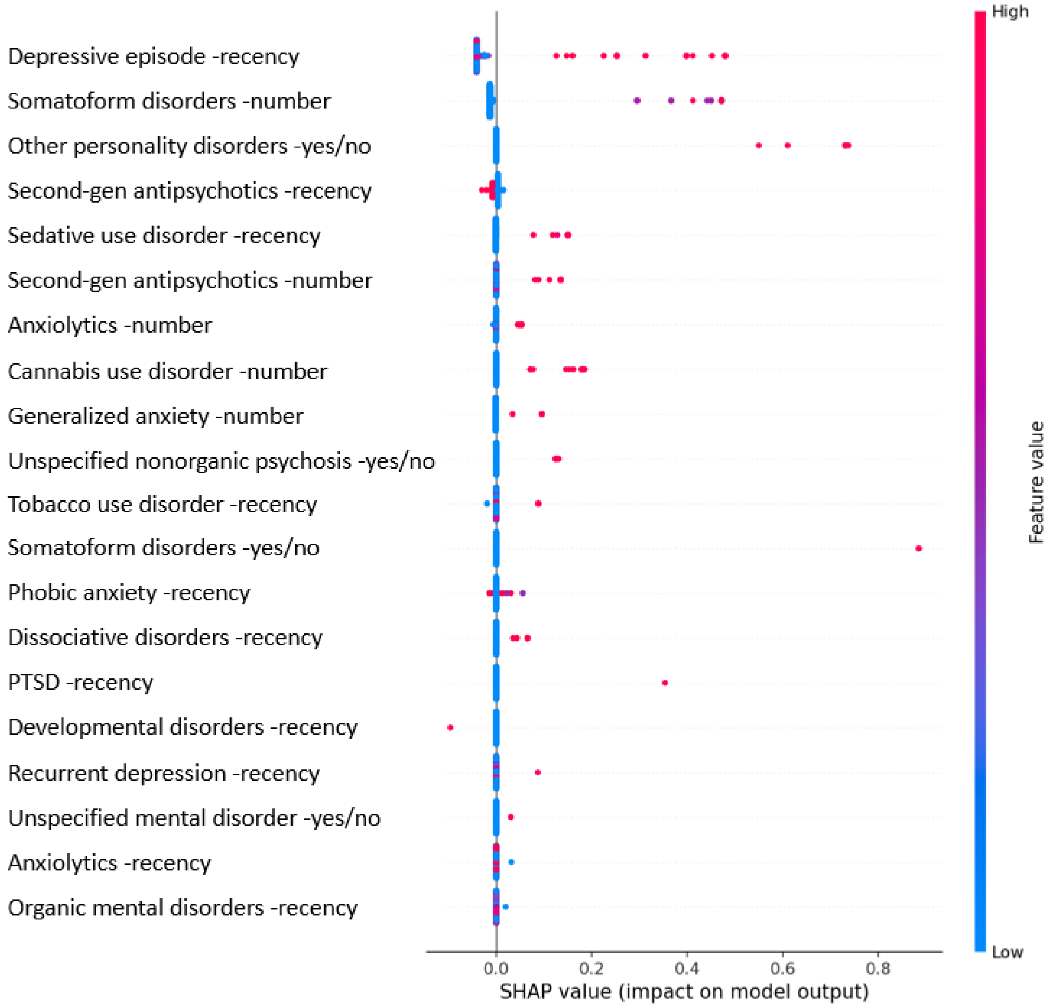


The colour of each point on the graph represents the value of the corresponding variable: pink indicates high values and blue indicates low values. The horizontal axis (x-axis) represents the SHAP value.
